# Mitigating Atherosclerotic Cardiovascular Disease Risk in Individuals with Elevated Lipoprotein(a): The Role of Modifiable Risk Factors

**DOI:** 10.1101/2025.05.22.25328169

**Authors:** Fu-Rong Li, Daniel Nyarko Hukportie

**Author notes:** Correspondence to: Fu-Rong Li School of Public Health and Emergency Management, Southern University of Science and Technology, Shenzhen, Guangdong, 518055, China.

## Abstract

**Importance:** Lipoprotein(a) [Lp(a)] is a primarily hereditary lipid linked to an increased risk of atherosclerotic cardiovascular disease (ASCVD). Currently, there are no pharmacological treatments specifically targeting elevated Lp(a) to reduce ASCVD risk.

**Objective:** To examine whether achieving optimal levels of modifiable risk factors could attenuate the ASCVD risk associated with elevated Lp(a).

**Design, Setting, and Participants:** Prospective cohort analysis of 289,186 ASCVD-free participants from the UK Biobank, including 57,813 individuals with elevated circulating Lp(a) and 231,373 individuals with non-elevated Lp(a) (control group). Findings were further validated in the ARIC study.

**Exposures:** Elevated Lp(a) was defined as either the highest ethnicity-/race-specific quintile of circulating Lp(a) levels or based on an Lp(a) polygenic risk score (PRS). Modifiable risk factor within target ranges included body mass index, diet quality, physical activity, smoking status, sleep duration, blood pressure, glycemic control, and low-density lipoprotein cholesterol (LDL-C).

**Main Outcomes and Measures:** The primary outcome was ASCVD. Cox proportional hazards models were used to estimate hazard ratios (HRs) and 95% confidence intervals (CIs).

**Results:** Over a median follow-up of 12.6 years, 12,592 ASCVD events were recorded. Among participants with elevated Lp(a), the risk of ASCVD progressively decreased as the number of modifiable risk factor within target ranges increased. Individuals with 7–8 risk factor within target ranges had a 24% lower risk of ASCVD (HR, 0.76; 95% CI, 0.70-0.82) compared with the control group. The most significant risk factors were smoking, systolic blood pressure, and LDL-C. The beneficial effects of controlling risk factors may be partly mediated through inflammatory pathways. Results were consistent when analyzed using the Lp(a) PRS in the UK Biobank and were confirmed in the ARIC study.

**Conclusions and Relevance:** Optimal management of modifiable risk factors may reduce the excess ASCVD risk associated with elevated Lp(a). These findings underscore the importance of comprehensive risk factor control in individuals with high Lp(a) levels.

## Introduction

Lipoprotein(a) [Lp(a)] has drawn special attention from health experts worldwide since its discovery in 1963(1). This lipid biomarker comprises of a covalently bound low-density lipoprotein (LDL)-like moiety and apolipoprotein(a). Mounting evidence has consistently demonstrated a causal role of elevated Lp(a) in the development of atherosclerotic cardiovascular disease (ASCVD)(2–3). Notably, on an equimolar basis, Lp(a) exhibits a greater atherogenic potential when compared to low-density lipoprotein cholesterol (LDL-C)(4).

Circulating Lp(a) levels are largely genetically determined and are predominantly influenced by genetic variants in the LPA gene(5). It has been estimated that approximately 1.4 billion individuals worldwide have elevated Lp(a) (≥50 mg/dL), resulting in a global prevalence ranging from 20% to 30%(6). These findings underscore the alarming prevalence of elevated Lp(a) among the general population. Unfortunately, however, there are no medications specifically approved for lowering ASCVD risk associated with elevated Lp(a), although some potential Lp(a)-lowering therapies are currently being evaluated in Phase II/III clinical trials(7). As such, interventions aimed at mitigating ASCVD risk in individuals with elevated Lp(a) primarily focus on addressing accompanying risk factors, such as lifestyle modifications and management of other cardiovascular risk factors like blood pressure, blood glucose, and lipid profiles(8).

Epidemiological evidence is limited regarding the extension of the beneficial effects of risk factor control on lowering ASCVD risks associated with elevated Lp(a). In the EPIC-Norfolk population study, researchers found ideal cardiovascular health to be associated with lower cardiovascular disease (CVD) risk, regardless of Lp(a) concentration or genotype(9). However, the relative significance of specific risk factors contributing to ASCVD risk, and the extent of risk reduction associated with optimal levels of a combination of behavioral or biological risk factors were not clearly elucidated, whiles only a single nucleotide polymorphism (SNP) was used in the genetic analysis. In addition, since risk factors that may contribute to the development of ASCVD are likely to differ between people from different regions of the world, further studies with diverse populations are warranted.

Leveraging on data from two large population-based cohorts from the United Kingdom (the UK Biobank study) and the United States (the Atherosclerosis Risk in Community [ARIC] study), we sought to investigate the impact of achieving optimal target ranges for modifiable risk factors on the Lp(a)-ASCVD relationship and the relative importance of risk factors for ASCVD risk among those with elevated Lp(a).

## Methods

The study protocol of the UK Biobank received ethical approval from the U.K. North West Multicenter Research Ethics Committee. The ARIC study underwent comprehensive evaluation and obtained approval from the National Heart, Lung, and Blood Institute (NHLBI) and the local institutional review boards affiliated with the participating clinical sites. Written informed consent was obtained from all participants during the enrollment process.

### Study Population

The study designs of the UK Biobank and ARIC have been previously described (10–11). Detailed descriptions of the two studies are presented in the **Supplementary materials**. We included UK Biobank and ARIC participants who were free of prevalent ASCVD and had information available regarding circulating Lp(a) levels, relevant covariates, fasting >8 h (applicable to ARIC only), and ASCVD events during follow-up. The final study sample utilized for the analysis of circulating Lp(a) consisted of 289,186 participants from UK Biobank and 11,524 participants from ARIC; as for the analyses involving Lp(a) polygenetic risk score (PRS) in the UK Biobank, we further excluded participants with missing genotype data, high relatedness or non-White/European ethnicity, leaving an analytic sample of 273,998 participants. Participants who were lost to follow-up through September 30, 2021, in UK Biobank and through December 31, 2017, in ARIC were censored in survival analyses.

### Circulating Lp(a), LPA PRS and risk factor definitions

Circulating Lp(a) measurement in the UK Biobank employed the Randox AU5800 platform (Randox Bioscience, UK) through immunoturbidimetric analysis, which is not influenced by isoform(12). In the ARIC study, circulating Lp(a) was measured by assessing its total protein composition, consisting of apolipoprotein(a) and apolipoprotein B. This assessment used a double-antibody ELISA technique specifically designed for detecting Lp(a)(13).

Genotyping array data obtained from participants of the UK Biobank were utilized to compute a previously established weighted PRS for Lp(a) levels. A prior investigation has shown that this Lp(a) PRS exhibited a comparable predictive capacity for ASCVD as circulating Lp(a) levels(14). **Supplementary Table S1** provides comprehensive information regarding the 43 SNPs included in the analysis. A weighted PRS was derived by employing these SNPs, where each variant was assigned a weight corresponding to its impact on circulating Lp(a) levels. The PRS for Lp(a) ranged from -140.76 to 388.40, with higher scores indicating higher levels of circulating Lp(a).

The determination of whether the risk factors fell within target ranges was largely established by referencing the cardiovascular health metrics recommended by the American Heart Association (AHA)(15). Accordingly, we selected five behavioral and three biological factors as the risk factors of interest in the UK Biobank. The behavioral risk factors included body mass index (BMI), diet quality, regular exercise, smoking, and sleep duration, while the biological risk factors include levels of blood pressure, glycated hemoglobin (HbA1c), and LDL-C. Due to the lack of data on sleep duration during the baseline visit of the ARIC study conducted between 1987 and 1989, this factor was not included in the analysis. Additionally, HbA1c measurements were not available in ARIC, and therefore fasting plasma glucose (FPG) levels were used as a substitute for glycemic control assessment. For methodological consistency, we did not adjust LDL-C values for Lp(a) to preserve clinically measured LDL-C, thereby better reflecting real-world clinical practice. The definitions of the risk factor within target ranges are provided in **Supplementary Tables S2-S3**.

### ASCVD Outcomes

For both cohorts, the primary outcome was ASCVD, which was a composite outcome of myocardial infarction (MI), peripheral arterial disease (PAD), ischemic stroke (IS), and CVD death. In the UK Biobank, the ascertainment of ASCVD relied upon the records of International Statistical Classification of Diseases and Related Health Problems (ICD) codes, in conjunction with Office of Population Censuses and Surveys Classification of Interventions and Procedures (OPCS) codes (**Supplementary Table S4**). Detailed descriptions of the methodologies employed in the ARIC study for the evaluation of incident fatal and nonfatal MI and IS have been previously elucidated(16–17). Events related to PAD were identified through hospitalizations where diagnostic codes corresponding to PAD (ICD-9: 440.2, 440.3, 440.4) or procedure codes indicative of leg revascularization (38.18, 39.25, 39.29, 39.50)(18).

### Statistical Analysis

Crude incidence rates were calculated according to the number of risk factor within target ranges and are presented as events per 1,000 person-y of observation. Bootstrap resampling was performed to estimate the 95% confidence intervals (CIs). Multivariable-adjusted Cox models were used to estimate the hazard ratios (HRs) and corresponding 95% CIs. All the models were adjusted for age, sex, ethnicity, Townsend deprivation index (TDI, for UK Biobank), family income (for ARIC), family history (FHx) of CVD, antihyperglycemic drug use, antihypertensive drug use, and statin use. For analyses regarding PRS, the first 10 principal components of ancestry were further adjusted for. No violation of the proportional hazard assumption was observed when examining Schoenfeld residuals through graphical diagnostics.

Considering the established racial variations in Lp(a) levels(19), we grouped the sample into ethnicity-/race-specific Lp(a) quintiles, and the first to fourth quintile was considered as non-elevated circulating Lp(a), and the fifth quintile as elevated circulating Lp(a). For analyses that were involved in PRS, we restricted the sample to the White only, and low Lp(a) genetic predisposition was defined as the first to fourth quintile of Lp(a) PRS, and high Lp(a) genetic predisposition as the highest quintile. For the main analyses, we estimated the risk of each outcome among participants with elevated Lp(a), according to the number of risk factor within target ranges, as compared with those with non-elevated Lp(a). Among participants with elevated Lp(a) in UK Biobank, the following categories of risk factor within target ranges were generated: 0-2, 3-4, 5-6 and 7-8. As for participants with elevated Lp(a) in ARIC, the following categories of risk factor within target ranges were retained in the analysis to include groups that were sufficiently large: 0-3, 4-5, and 6-7. To evaluate the relative importance of the risk factors among participants with elevated Lp(a), an analysis was conducted to ascertain their respective contributions in predicting the outcomes. This assessment was performed by examining the R^2^ values derived from the models(20).

In the exploratory analysis, we further adjusted for hypersensitive C-reactive protein (hs-CRP), white blood cell (WBC) count, and platelet count, in an attempt to examine if the inflammatory pathways and thrombotic pathways may potentially mediate the protective effects of multiple risk factors on ASCVD risk. We also examined whether the associations investigated differed by age (<50 y, 50-59 y, and ≥60 y), sex and 10-y ASCVD risk categories [10% based on QRISK3 for UK Biobank(21), and 7.5% based on PCE for ARIC(22)]. Interactions were tested by adding a multiplicative term to the Cox models.

Several sensitivity analyses were also conducted: 1) redefining the circulating elevated Lp(a) as Lp(a) levels of ≥120 nmol/L (≈50 mg/dL) for UK Biobank and ≥50 mg/dL for ARIC; 2) redefining the control group as those with non-elevated circulating Lp(a) or low Lp(a) PRS but with median risk factors within target ranges, namely 4 risk factor within target ranges in UK Biobank, and 3-4 risk factor within target ranges in ARIC; 3) redefining the control group as those with lowest quintile of circulating Lp(a) or Lp(a) PRS; and 4) exclusion of those who developed ASCVD within 5 years of follow-up after the baseline assessment.

## Results

### Baseline Characteristics

The study population of UK Biobank included 57,813 individuals with elevated circulating Lp(a) (i.e., quintile 5) and 231,373 control with non-elevated circulating Lp(a) (i.e., quintile 1-4). In addition, 54,753 participants had high Lp(a) PRS and 219,245 had low Lp(a) PRS. In the ARIC study, we included 2,290 participants with elevated circulating Lp(a) and 9,234 control with non-elevated circulating Lp(a) (**Supplementary Figure S1**).

Among participants with elevated circulating Lp(a) in the UK Biobank, an increasing number of risk factor within target ranges was associated with better risk profiles, including lower levels of hs-CRP, WBC count and platelet count, younger age, lower TDI and more likely to be female sex but less likely to have a FHx of CVD and were not taking antihyperglycemic, antihypertensive, and lipid-lowering medications (**Table 1**). Similar patterns were also observed when stratifying the UK Biobank participants by Lp(a) PRS or stratifying the ARIC study by circulating Lp(a) (**Supplementary Tables S5-S6**).

**Table 1.**
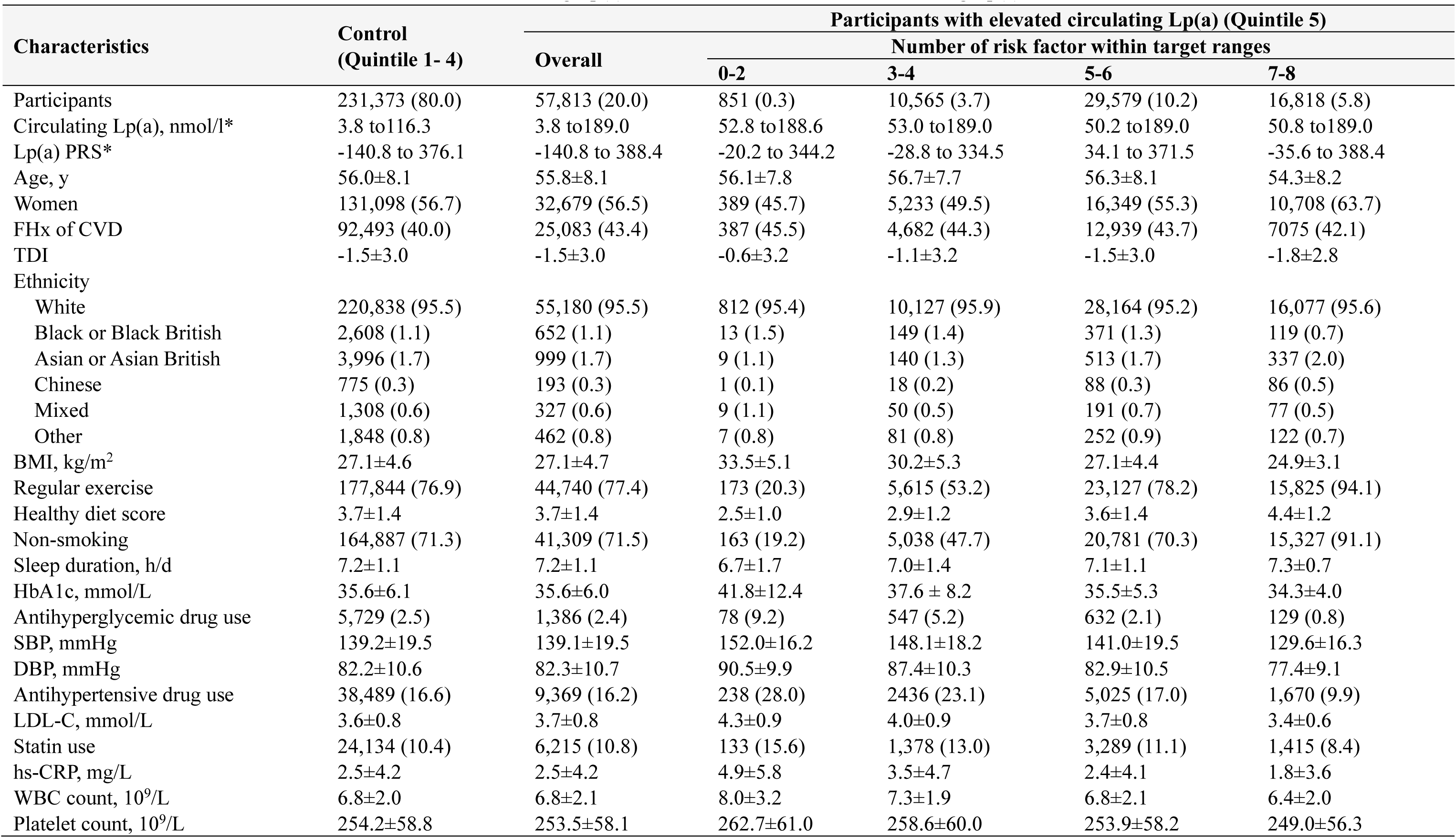
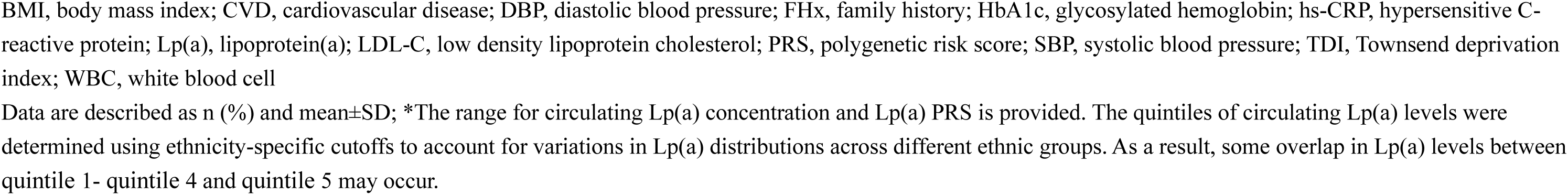
Characteristics of individuals with non-elevated circulating Lp(a) and individuals with elevated circulating Lp(a) from the UK Biobank.

| Characteristics | Control<br>(Quintile 1- 4) | Participants with elevated circulating Lp(a) (Quintile 5) |  |  |  |  |
| --- | --- | --- | --- | --- | --- | --- |
|  |  | Overall | Number of risk factor within target ranges |  |  |  |
|  |  |  | 0-2 | 3-4 | 5-6 | 7-8 |
| Participants | 231,373 (80.0) | 57,813 (20.0) | 851 (0.3) | 10,565 (3.7) | 29,579 (10.2) | 16,818 (5.8) |
| Circulating Lp(a), nmol/l* | 3.8 to116.3 | 3.8 to189.0 | 52.8 to188.6 | 53.0 to189.0 | 50.2 to189.0 | 50.8 to189.0 |
| Lp(a) PRS* | -140.8 to 376.1 | -140.8 to 388.4 | -20.2 to 344.2 | -28.8 to 334.5 | 34.1 to 371.5 | -35.6 to 388.4 |
| Age, y | 56.0±8.1 | 55.8±8.1 | 56.1±7.8 | 56.7±7.7 | 56.3±8.1 | 54.3±8.2 |
| Women | 131,098 (56.7) | 32,679 (56.5) | 389 (45.7) | 5,233 (49.5) | 16,349 (55.3) | 10,708 (63.7) |
| FHx of CVD | 92,493 (40.0) | 25,083 (43.4) | 387 (45.5) | 4,682 (44.3) | 12,939 (43.7) | 7075 (42.1) |
| TDI | -1.5±3.0 | -1.5±3.0 | -0.6±3.2 | -1.1±3.2 | -1.5±3.0 | -1.8±2.8 |
| Ethnicity |  |  |  |  |  |  |
| White | 220,838 (95.5) | 55,180 (95.5) | 812 (95.4) | 10,127 (95.9) | 28,164 (95.2) | 16,077 (95.6) |
| Black or Black British | 2,608 (1.1) | 652 (1.1) | 13 (1.5) | 149 (1.4) | 371 (1.3) | 119 (0.7) |
| Asian or Asian British | 3,996 (1.7) | 999 (1.7) | 9 (1.1) | 140 (1.3) | 513 (1.7) | 337 (2.0) |
| Chinese | 775 (0.3) | 193 (0.3) | 1 (0.1) | 18 (0.2) | 88 (0.3) | 86 (0.5) |
| Mixed | 1,308 (0.6) | 327 (0.6) | 9 (1.1) | 50 (0.5) | 191 (0.7) | 77 (0.5) |
| Other | 1,848 (0.8) | 462 (0.8) | 7 (0.8) | 81 (0.8) | 252 (0.9) | 122 (0.7) |
| BMI, kg/m<sup>2</sup> | 27.1±4.6 | 27.1±4.7 | 33.5±5.1 | 30.2±5.3 | 27.1±4.4 | 24.9±3.1 |
| Regular exercise | 177,844 (76.9) | 44,740 (77.4) | 173 (20.3) | 5,615 (53.2) | 23,127 (78.2) | 15,825 (94.1) |
| Healthy diet score | 3.7±1.4 | 3.7±1.4 | 2.5±1.0 | 2.9±1.2 | 3.6±1.4 | 4.4±1.2 |
| Non-smoking | 164,887 (71.3) | 41,309 (71.5) | 163 (19.2) | 5,038 (47.7) | 20,781 (70.3) | 15,327 (91.1) |
| Sleep duration, h/d | 7.2±1.1 | 7.2±1.1 | 6.7±1.7 | 7.0±1.4 | 7.1±1.1 | 7.3±0.7 |
| HbA1c, mmol/L | 35.6±6.1 | 35.6±6.0 | 41.8±12.4 | 37.6 ± 8.2 | 35.5±5.3 | 34.3±4.0 |
| Antihyperglycemic drug use | 5,729 (2.5) | 1,386 (2.4) | 78 (9.2) | 547 (5.2) | 632 (2.1) | 129 (0.8) |
| SBP, mmHg | 139.2±19.5 | 139.1±19.5 | 152.0±16.2 | 148.1±18.2 | 141.0±19.5 | 129.6±16.3 |
| DBP, mmHg | 82.2±10.6 | 82.3±10.7 | 90.5±9.9 | 87.4±10.3 | 82.9±10.5 | 77.4±9.1 |
| Antihypertensive drug use | 38,489 (16.6) | 9,369 (16.2) | 238 (28.0) | 2436 (23.1) | 5,025 (17.0) | 1,670 (9.9) |
| LDL-C, mmol/L | 3.6±0.8 | 3.7±0.8 | 4.3±0.9 | 4.0±0.9 | 3.7±0.8 | 3.4±0.6 |
| Statin use | 24,134 (10.4) | 6,215 (10.8) | 133 (15.6) | 1,378 (13.0) | 3,289 (11.1) | 1,415 (8.4) |
| hs-CRP, mg/L | 2.5±4.2 | 2.5±4.2 | 4.9±5.8 | 3.5±4.7 | 2.4±4.1 | 1.8±3.6 |
| WBC count, 10<sup>9</sup>/L | 6.8±2.0 | 6.8±2.1 | 8.0±3.2 | 7.3±1.9 | 6.8±2.1 | 6.4±2.0 |
| Platelet count, 10<sup>9</sup>/L | 254.2±58.8 | 253.5±58.1 | 262.7±61.0 | 258.6±60.0 | 253.9±58.2 | 249.0±56.3 |

BMI, body mass index; CVD, cardiovascular disease; DBP, diastolic blood pressure; FHx, family history; HbA1c, glycosylated hemoglobin; hs-CRP, hypersensitive C-reactive protein; Lp(a), lipoprotein(a); LDL-C, low density lipoprotein cholesterol; PRS, polygenetic risk score; SBP, systolic blood pressure; TDI, Townsend deprivation index; WBC, white blood cell
Data are described as n (%) and mean ± SD; \*The range for circulating Lp(a) concentration and Lp(a) PRS is provided. The quintiles of circulating Lp(a) levels were determined using ethnicity-specific cutoffs to account for variations in Lp(a) distributions across different ethnic groups. As a result, some overlap in Lp(a) levels between quintile 1- quintile 4 and quintile 5 may occur.

### Lp(a), risk factors and risk of ASCVD

In the UK Biobank, after a median follow-up of approximately 12 years, 3,725 of individuals with elevated circulating Lp(a) and 3,504 of individuals with high Lp(a) PRS developed ASCVD. In the ARIC study, 700 of individuals with elevated circulating Lp(a) developed ASCVD during a median follow-up of 26.1 years. Elevated Lp(a), whether in terms of circulating levels or PRS, were both associated with higher risks of all ASCVD outcomes incidence, with HRs ranging from 1.08 to 1.32, although the excess risk of IS associated with high Lp(a) PRS in the UK Biobank was not statistically significant (**Supplementary Table S7**). In most cases, risk factor within target ranges were associated with lower risks of all ASCVD outcomes among participants with elevated circulating Lp(a) or high Lp(a) genetic predisposition (**Supplementary Tables S8-S9**). **Figure 1** shows the predictors with the apparent greatest importance in relation to ASCVD. The three strongest predictors for ASCVD were smoking, SBP and LDL-C in both cohorts. Although the strongest predictors for the remaining specific ASCVD outcomes varied, smoking and SBP appeared to be the most common top three predictors in both cohorts (**Supplementary Figures S2-S4**). When jointly considering the number of risk factor within target ranges, we found significant inverse associations between the degree of optimal risk factor levels and the risks of all ASCVD outcomes (all *P*-trend <0.001 for both cohorts); specifically, each additional optimal risk factor was linked to a 21% (HR= 0.79) to 26% (HR= 0.74) lower risks of ASCVD across the two cohorts (**Supplementary Tables S10-S11**).

**Figure 1.**
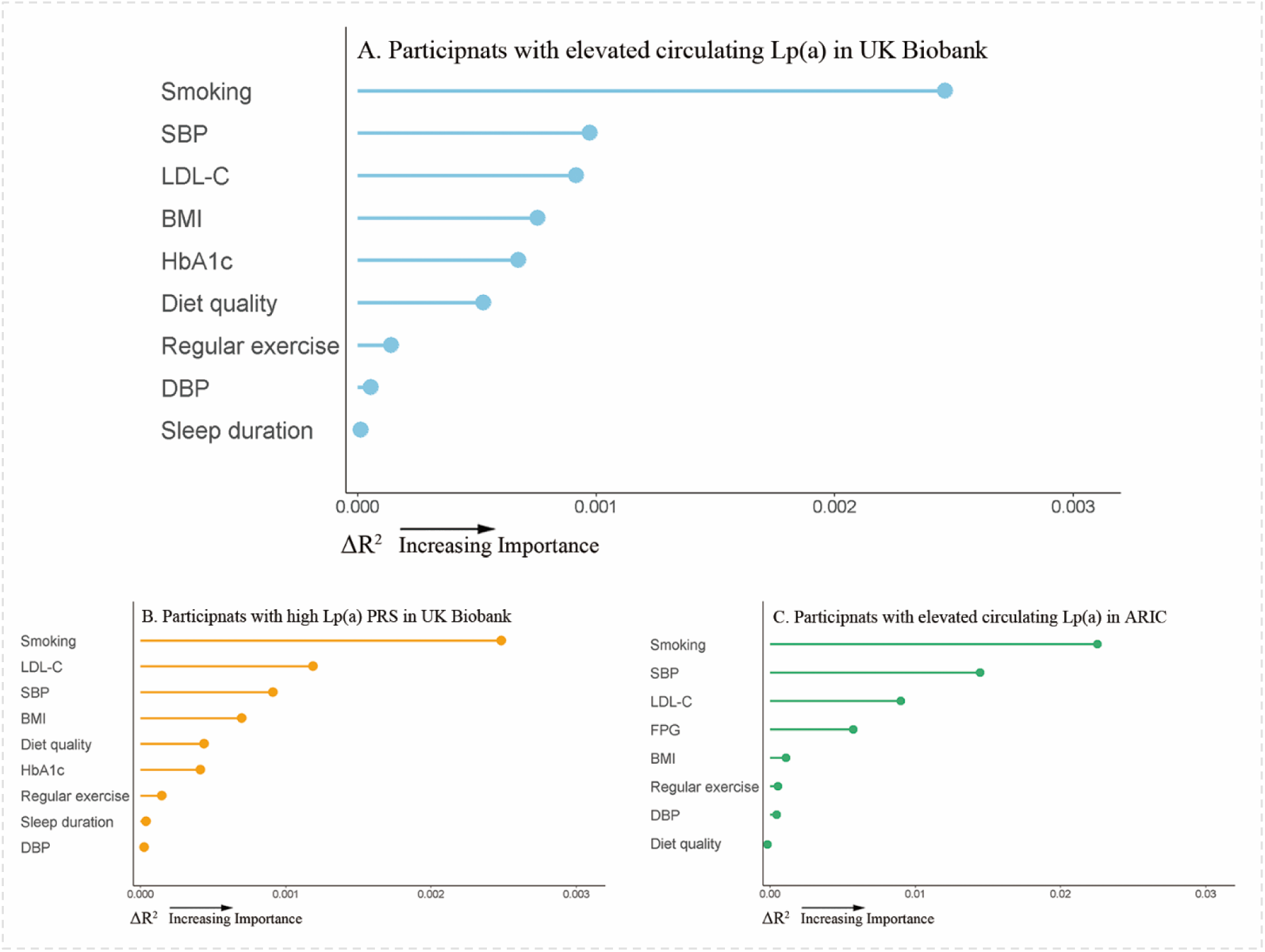
Relative importance of risk factors for predicting ASCVD in UK Biobank and ARIC. The ΔR^2^ value for each risk factor was computed by subtracting the R^2^ value of the Cox model that did not include the specific risk factor from the R^2^ value generated by the Cox model that included all the risk factors. For participants with high Lp(a) PRS, the first 10 principal components of ancestry was further adjusted for, and the analysis was conducted among the White. ARIC, Atherosclerosis Risk in Communities; BMI, body mass index; DBP, diastolic blood pressure; FPG, fasting plasma glucose; HbA1c, glycosylated hemoglobin; LDL-C, low density lipoprotein cholesterol; Lp(a), lipoprotein(a); PRS, polygenetic risk score; SBP, systolic blood pressure

### ASCVD risk according to the number of risk factor within target ranges

**Supplementary Tables S12-S15** show the incidence rates and absolute rate differences for overall and specific ASCVD outcomes for individuals with elevated Lp(a) and according to the number of risk factor within target ranges. Generally, incidence rates of all ASCVD outcomes decreased stepwise with an increasing number of risk factor within target ranges.

Overall, we found that those with elevated Lp(a) but had highest number of risk factor within target ranges even had lower ASCVD risk in comparison to those with non-elevated Lp(a). In UK Biobank, HRs (95% CIs) for those with elevated circulating Lp(a) and with 0-2, 3-4, 5-6, and 7-8 risk factor within the target ranges were 2.38 (2.00, 2.84), 1.74 (1.64, 1.85), 1.17 (1.12, 1.23), and 0.76 (0.70, 0.82), respectively, as compared with individuals with non-elevated circulating Lp(a); similarly when considering Lp(a) levels in terms of PRS. In the ARIC study, we also found a consistent decrease in the excess risk of ASCVD with a higher number of risk factor within the target ranges among those with elevated circulating Lp(a), compared with participants with non-elevated circulating Lp(a); the HRs (95% CIs) for 0-3, 4-5, and 6-7 risk factor within target ranges were respectively 1.94 (1.64, 2.29), 1.37 (1.23, 1.53), and 0.80 (0.69, 0.94) (**Figure 2**). Similar patterns of associations were also noted for each specific ASCVD outcomes (**Figure 3**).

**Figure 2.**
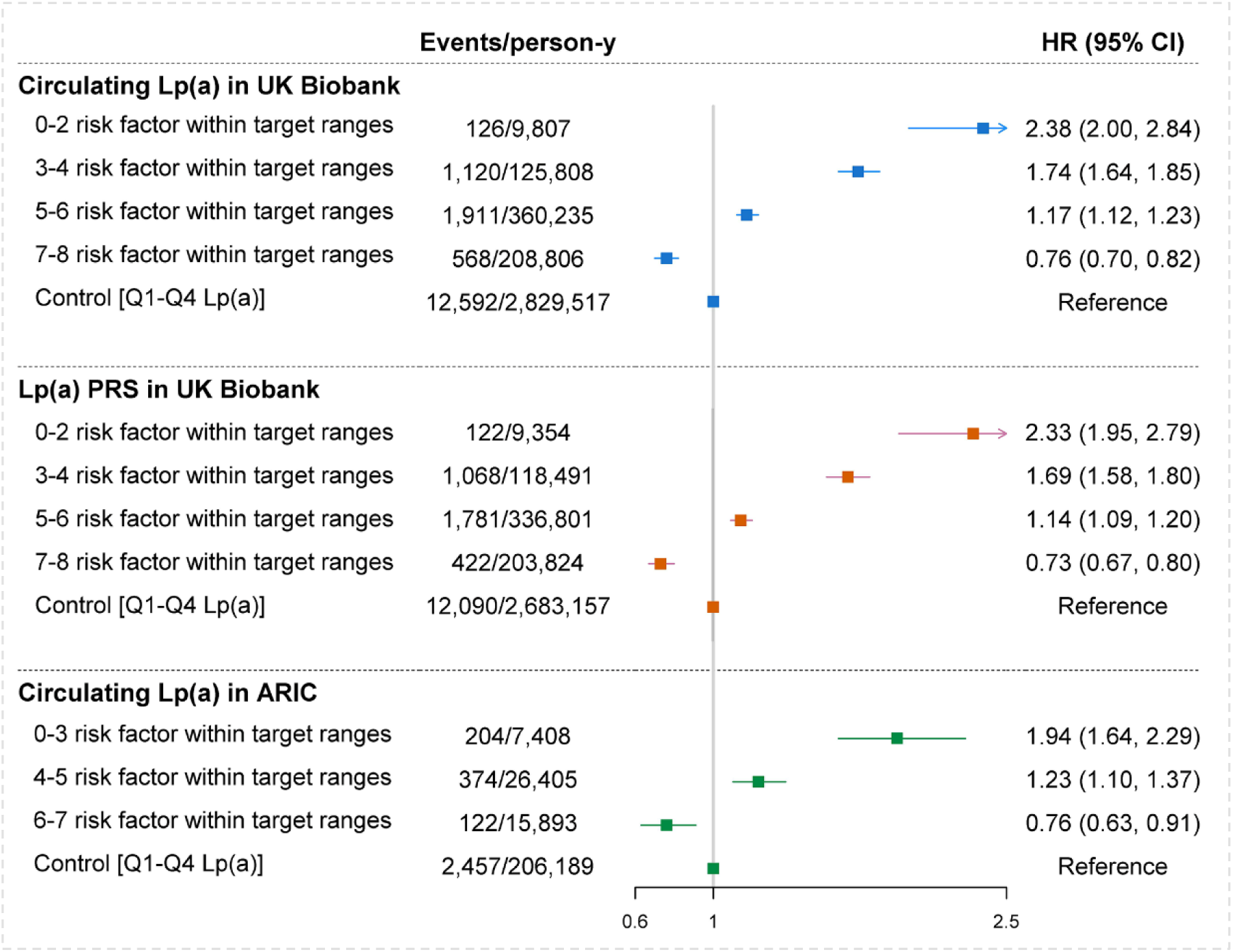
HRs for ASCVD according to the number of risk factors at target among participants with elevated circulating Lp(a) or high Lp(a) PRS compared with those with non-elevated circulating Lp(a) or low Lp(a) PRS in UK Biobank and ARIC. For circulating Lp(a) in UK Biobank, the model was adjusted for age, sex, ethnicity, TDI, FHx of CVD, antihyperglycemic drug use, antihypertensive drug use, and statin use. For Lp(a) PRS in UK Biobank, the analyses were conducted among the White only. The model was adjusted for the covariates that were used for circulating Lp(a) and further adjusted for the first 10 principal components of ancestry. For circulating Lp(a) in ARIC, the model was adjusted for age, sex, ethnicity, family income, FHx of CVD, antihyperglycemic drug use, antihypertensive drug use, and statin use. ARIC, Atherosclerosis Risk in Communities; CI, confidence interval; HR, hazard ratio; Lp(a), lipoprotein(a)

**Figure 3.**
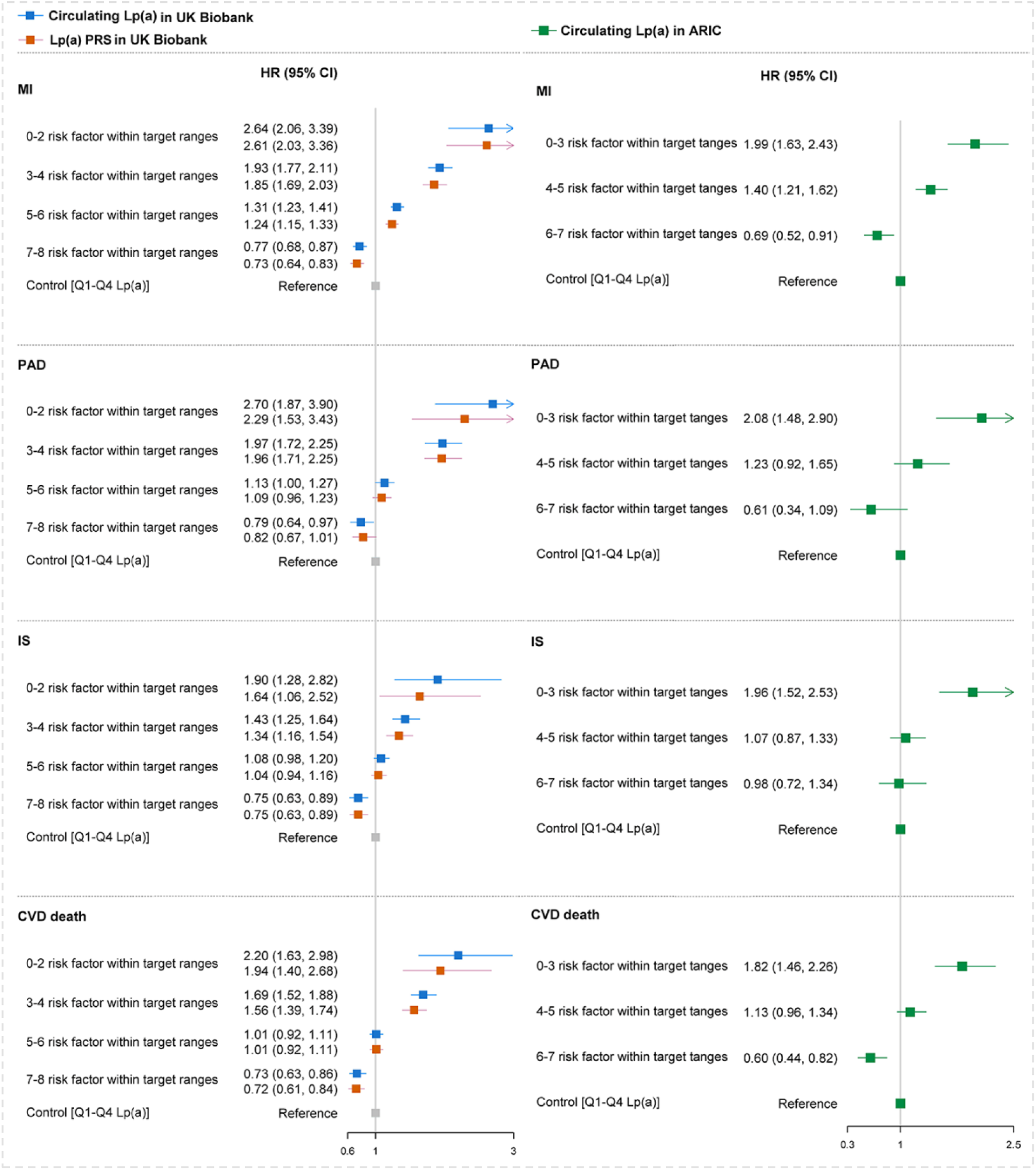
HRs for individual ASCVD outcomes according to the number of risk factors at target among participants with elevated circulating Lp(a) or high Lp(a) PRS compared with those with non-elevated circulating Lp(a) or low Lp(a) PRS in UK Biobank and ARIC. For circulating Lp(a) in UK Biobank, the model was adjusted for age, sex, ethnicity, TDI, FHx of CVD, antihyperglycemic drug use, antihypertensive drug use, and statin use. For Lp(a) PRS in UK Biobank, the analyses were conducted among the White only. The model was adjusted for the covariates that were used for circulating Lp(a) and further adjusted for the first 10 principal components of ancestry. For circulating Lp(a) in ARIC, the model was adjusted for age, sex, ethnicity, family income, FHx of CVD, antihyperglycemic drug use, antihypertensive drug use, and statin use. Abbreviations: CVD, cardiovascular disease; Lp(a), lipoprotein(a); MI, myocardial infarction; PAD, peripheral arterial disease; PRS, polygenetic risk score; IS, ischemic stroke.

### Exploratory analysis and sensitivity analysis

In both cohorts, the associations observed in the main analyses appeared to be attenuated after adjusting for inflammatory biomarkers (i.e., hs-CRP and/or WBC count), but not thrombotic biomarker (i.e., platelet count) (**Table 2**). There were no statistically significant interactions for age or sex regarding main analyses in both UK Biobank and the ARIC Study (all *P*-interactions >0.200) (**Supplementary Tables S16-S18**). Despite evidence of interaction, having risk factors within target ranges showed no harmful effect on ASCVD risk, regardless of high or low estimated 10-y ASCVD risk. (**Supplementary Tables S19**). The main results were not materially changed in a series of sensitivity analyses in both cohorts (**Supplementary Tables S20-S22**).

**Table 2.**
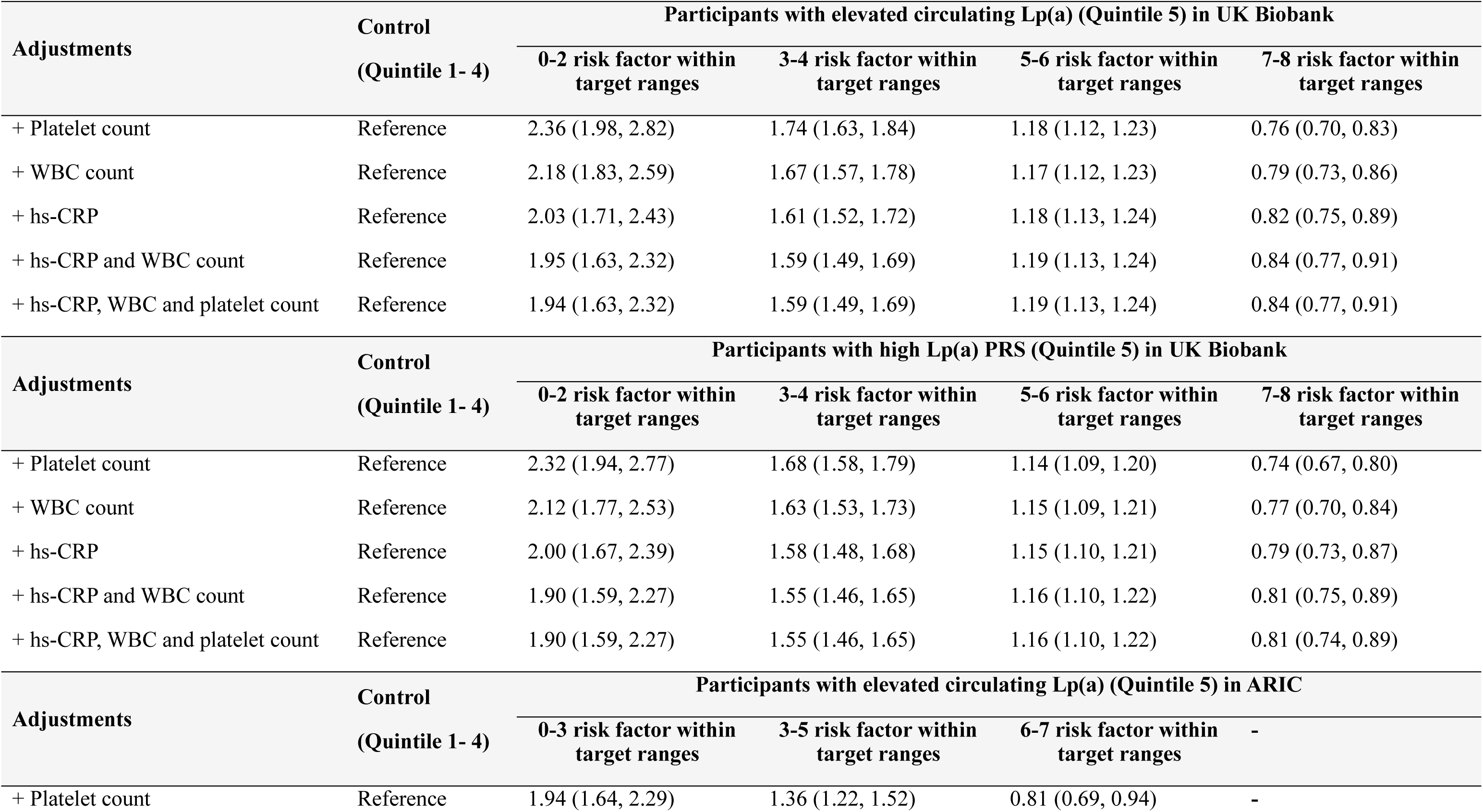
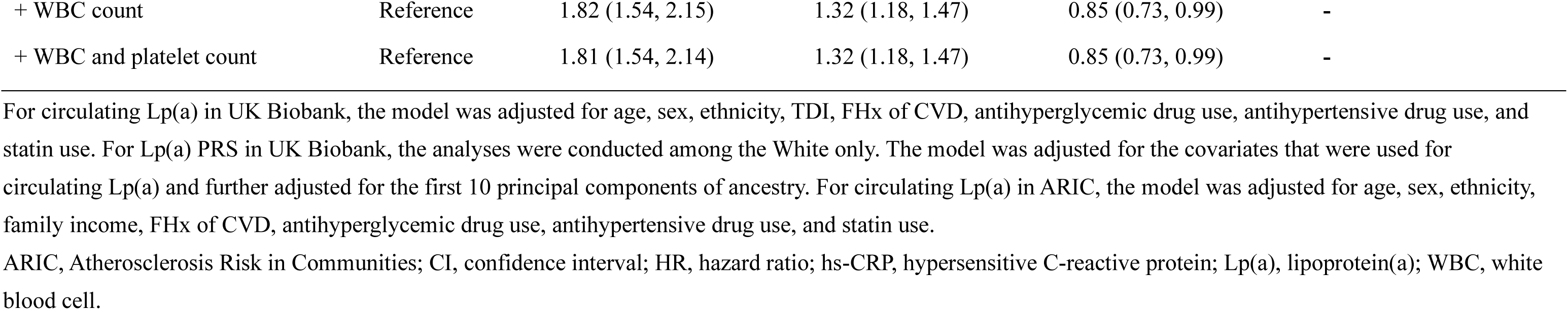
Further adjustments for inflammatory biomarkers and thrombotic biomarker*.

## Discussion

In the present study, we examined the impact of attaining modifiable behavioral and biological risk factor within target ranges on the relationship between circulating Lp(a) or Lp(a) PRS and ASCVD in two large population-based cohorts from the UK and the US. We found the hazardous link between elevated Lp(a), whether in terms of circulating levels or genetic predisposition, and risks of all ASCVD outcomes incidence to decrease in a stepwise fashion with an increasing number of predefined risk factor within the target ranges. Notably, having optimal risk factor levels appeared to eliminate ASCVD risk and even offered more favorable ASCVD outcomes among those with elevated Lp(a) compared to those with non-elevated Lp(a) or low genetic predisposition. These protective associations were largely similar across age, sex and estimated 10-y ASCVD risk profiles. Smoking, SBP, and LDL-C appeared to be the top 3 most important risk factors that should be principally targeted to mitigate ASCVD risk among individuals with elevated Lp(a).

Recent scientific statements from atherosclerosis societies called for intensified lifestyle and joint CVD risk factor intervention in the presence of elevated Lp(a) (8, 23–24). Currently, however, randomized controlled trials examining the modifying effect of joint behavioral or biological CVD risk factor control on Lp(a)-ASCVD link is lacking, although a recent study of U.S. cohorts found that modifiable risk factors have only a modest effect on the associations of Lp(a) with MI and stroke(25). In a recent registry-based study, Shiyovich *et al.* assessed the risk of acute MI linked to elevated Lp(a) relative to modifiable risk factors, but did not examine the potential mitigating effects of these factors(26). In an observational study of approximately 14,000 individuals enrolled in the EPIC-Norfolk (mean follow-up time: 11.5 years), Perrot *et al*. found that having ideal cardiovascular health metrics appear to have a 67% and 76% lower risk of developing a composite outcome of coronary heart disease and stroke among those with elevated Lp(a) and Lp(a)-raising allele, respectively, in comparison with those with poor cardiovascular health and non-elevated Lp(a)(9), pointing to the valuable application of attaining a higher number of risk factor within target ranges in individuals with elevated Lp(a). In the current study, we provide new epidemiological evidence using data on both circulating Lp(a) levels and 43 SNPs derived Lp(a) PRS from two large population-based prospective cohort studies in the UK and the US. Our findings added to this literature by showing that having optimal levels of composite behavioral or biological risk factors in individuals with elevated Lp(a) levels or high genetic predisposition would decrease or even eliminate their risk of overall and specific ASCVD outcomes (i.e., MI, PAD, IS and CVD death).

We compared the relative importance of individual risk factors in predicting ASCVD among those with elevated Lp(a), and found the three strongest predictors for the composite ASCVD outcome to be smoking, SBP, and LDL-C in both study cohorts; an observation which was largely similar for specific ASCVD outcomes. This suggests that the ASCVD-associated risk due to elevated Lp(a) could be ameliorated, to a large degree, with the optimal control of these potent ASCVD-determining risk factors (i.e., smoking, SBP, and LDL-C). However, it is worth mentioning that, while having optimal levels of LDL-C appears to attenuate ASCVD risk associated with elevated Lp(a), residual ASCVD risk remained among individuals with elevated Lp(a)(27). This suggests that even when LDL-C levels are within the target range, only partial elimination of the ASCVD risk is achieved. Additionally, it has been observed that statins, commonly prescribed for lipid management, may contrarily elevate Lp(a) levels(28). Hence, we propose that the simultaneous control of all behavioral and biological risk factors, along with the management of LDL-C, will lead to a powerful “synergistic effect” that can effectively mitigate the excess ASCVD risk associated with elevated Lp(a).

Current expert consensus statement recommends personalized management approaches for individuals with elevated Lp(a) based on their risk profiles(8). Previous studies have suggested that younger individuals and women often exhibit a higher ASCVD risk in the presence of specific risk factors compared to older individuals and men(29–31). Additionally, a study from the UK Biobank found that elevated Lp(a) confers a greater risk of ASCVD in individuals with low-risk phenotypes compared to those with high-risk phenotypes(32). In our study, however, we found that across age, sex and 10-y ASCVD risk strata, individuals with the highest number of risk factor within target ranges consistently exhibited a lower ASCVD risk. This suggests that individuals with elevated Lp(a) should be equally targeted for aggressive ASCVD preventive strategies, regardless of age, sex and estimated ASCVD risk profiles.

The pro-inflammatory properties of Lp(a) are widely accepted as one of the major culprit driving the Lp(a)-ASCVD relationships(8, 33–34). Our analysis showed that the Lp(a)-ASCVD associations were attenuated after adjusting for inflammatory biomarkers, suggesting that the beneficial effects of controlling risk factors are likely to be mediated, at least in part, through inflammatory pathways. This highlights the significance of inflammation as a key underlying mechanism in the observed effects of risk factor control on ASCVD risk. Indeed, while achieving control of ASCVD-related risk factors may have a limited direct effect on circulating Lp(a) levels(24, 35), reaching and maintaining optimal levels of ASCVD-related risk factors may neutralize the pro-inflammatory activities of Lp(a). For instance, by achieving non-smoking target, the smoke-induced inflammatory reactions including the release of a host of reactive oxygen- and nitrogen species are eliminated, reducing the overall burden of inflammation(36–37). Achieving optimal blood pressure control helps to reduce oxidative stress, decrease the release of pro-inflammatory molecules and restore endothelial function(38). Also, physical activity(39), healthy diet habit(40), ideal sleep quality(41) and body weight(42), ideal glycemic control(43), and reduction in LDL-C levels(44) have all been associated with the ability to counteract inflammation, which is in contrast to the pro-inflammatory effects associated with elevated Lp(a).

Our study has several strengths, including a large sample size with participants from Europe and America, a prospective study design with long-term follow-up, and a comprehensive assessment of various ASCVD subtypes. However, the study findings need to be interpreted in light of some potential limitations. First, potential changes in risk factors over the study period were not taken into account, hence future studies may be needed to examine the impact of changes in risk factors on the Lp(a)-ASCVD relationship. Second, the subjective collection of data on healthy lifestyle information such as diet metrics and physical activity may be prone to recall bias. Third, the assay used in the ARIC study predated current lipoprotein(a) assays and could be influenced by changes in apo(a) isoforms due to variations in the number of Kringle 4 type 2 repeats(45). Nevertheless, the results from the UK Biobank cohort, which used a more contemporary Lp(a) assay, confirmed the finding from the ARIC cohort. Lastly, while our analysis indicates a stronger joint association between targeted risk factors and ASCVD risk among individuals with a low 10-y ASCVD risk, caution is warranted in interpreting these findings, as these factors are already part of ASCVD risk estimation, thus further studies are needed to validate and expand upon these findings.

In conclusion, while the elevated circulating Lp(a) level or high genetic predisposition to elevated Lp(a) leads to a higher risk of ASCVD, having optimal levels of behavioral and biological risk factors attenuates or even eliminates this excess risk. Hence, in the absence of medications approved to treat elevated Lp(a), achieving optimal behavioral and biological risk factor targets should be emphasized through intensive public health/medical programs to effectively reduce the risk of developing ASCVD in individuals with elevated Lp(a).

## Data Availability

All data produced in the present study are available upon reasonable request to the authors

https://zenodo.org/records/15491004

## Acknowledgements

We are grateful to the UK Biobank and AIRC participants. The research involving the UK Biobank was conducted under Application Number 60009.

## Author contributions

Fu-Rong Li and Fengchao Liang directed the study. Fu-Rong Li analyzed the data and wrote the manuscript. Dongfeng Gu, Daniel Nyarko Hukportie, Xia Li, Cheng Jin, Chunbao Mo, Jiangshui Wang, Xianbo Wu contributed to the discussion and reviewed/edited the manuscript. All authors contributed to the article and approved the submitted version.

