## Supplementary Table S1-S22; Supplementary Figure S1-S4 for "Mitigating Atherosclerotic Cardiovascular Disease Risk in Individuals with Elevated Lipoprotein(a): The Role of Modifiable Risk Factors"

**Supplementary Materials**

**Catalogue**

**Brief introduction of the UK Biobank study**

The UK Biobank is an ongoing prospective cohort comprising approximately 0.5 million individuals aged between 40 and 69 years. The cohort was recruited from 22 centers located across England, Wales, and Scotland during the period of 2006 to 2010 (1). During enrollment, the UK Biobank collected a wide range of measurements and extensive information on sociodemographic factors, physical measurements, lifestyle behaviors, and prevalent medical conditions. Furthermore, as part of the data collection, biological samples were also obtained by trained nurses for subsequent laboratory assays. ASCVD events and their incident date were ascertained by linking the study participants' data to the health episode statistics for individuals from England, and Wales and the Scottish Morbidity Records for those from Scotland. Information regarding the cause and date of death was obtained by linking the data to the death registries maintained by the National Health Service (NHS) Information Centre for participants from England and Wales, and the NHS Central Register Scotland for participants from Scotland.

Data on baseline characteristics were collected with a touch-screen questionnaire. Prevalent cardiovascular disease (CVD), including myocardial infarction (MI), stroke, and peripheral arterial disease (PAD) was identified by self-reported and inpatient records. In our analysis, we considered various factors including sociodemographic information (age, sex, ethnicity, and Townsend deprivation index [TDI]), lifestyle behaviors (smoking, diet quality, regular exercise, and sleep duration), as well as self-reported use of antihypertensive drugs, antihyperglycemic drugs, and statins. A non-stretchable tape was used to measure height, and the Tanita BC‐418 MA body analyser was used to measure weight (2). Body mass index (BMI) was calculated as weight in kilograms divided by height in metres squared (kg/m^2^). Usual dietary information was obtained using a touchscreen food-frequency questionnaire. This questionnaire included inquiries about the average intake of major foods or food groups over the past year. Following a similar approach as a previous study, a healthy diet was defined based on the consumption of seven commonly eaten food groups or dietary habits. This definition included higher consumption of fruit and vegetables, whole grains, and fish, lower consumption of red meat, processed meat, and refined grains, as well as avoiding the addition of salt to food (3-4). Those who reported a history of heart disease among their parents were defined as having a family history (FHx) of CVD (5). Two blood pressure (BP) measurements were taken while the participant was seated after 5 minutes of rest using an appropriate cuff and an Omron HEM-7015IT digital BP monitor. Mean systolic blood pressure (SBP) and diastolic blood pressure (DBP) values were calculated from 2 automated or 2 manual blood pressure measurements (6). Glycated hemoglobin (HbA1c) was measured by HPLC analysis on a Bio-Rad VARIANT II Turbo; this biomarker was used to reflect glycemic control because the blood samples were largely non-fasting in the UK Biobank and HbA1c is relatively stable within 2-3 months (7). Direct low-density lipoprotein cholesterol (LDL-C) was measured by enzymatic protective selection analysis on a Beckman Coulter AU5800 (8).

**Brief introduction of the Atherosclerosis Risk in Communities (ARIC) Study**

The Atherosclerosis Risk in Communities (ARIC) study is a prospective study involving approximately 15,000 individuals aged 45-64 years. The participants were recruited during the period from 1987 to 1989 (visit 1), hailing from four distinct communities situated across different regions of the United States, namely Minneapolis, Minnesota; Washington County, Maryland; Forsyth County, North Carolina; and Jackson, Mississippi (9). For most of the suspected cardiovascular events, thorough abstraction of medical records was conducted. Adjudication of ASCVD events was performed by an independent endpoints committee, ensuring the blinding of cardiovascular risk factor data to maintain objectivity and minimize potential biases. Mortality data were identified through rigorous methodologies, including utilization of the National Death Index and regular active surveillance, which involved semi-annual telephone contact with participants to evaluate the health status.

Prevalent CVD, including conditions such as coronary heart disease, stroke, and PAD, was assessed through a combination of self-reported information and inpatient records. Age, sex, ethnicity, family income, FHx of CVD, smoking status, physical activity, diet quality, medical history, and medication use were obtained through questionnaires administered to the participants. BMI was calculated as weight (in kilograms) divided by height (in meters)-squared. Family income was categorized as <$25,000, $25,000−$49,999, or ≥$50,000 per year. The definition of FHx of CVD was established when either the father or the mother had a history of CVD. The ARIC study used the Baecke Physical Activity questionnaire to assess usual sport, exercise and nonsport leisure. A semicontinuous index was calculated for each type of activity, ranging from 1 (low) to 5 (high). For example, the sports index was derived by considering factors such as the average yearly frequency, weekly duration, and intensity (low, medium, or high) of various sports or exercises. Dietary intake was evaluated utilizing a 66-item Food Frequency Questionnaire (FFQ), which was administered through interviewer-led assessments. The FFQ was a modified version of the 61-item Willett questionnaire and had undergone validation within the ARIC cohort (10-11). During Visit 1 (1987-1989), trained interviewers collected data from participants by asking them to report the average frequency of consuming specific food items, considering a given portion size, over the past year. Total energy and nutrient intake were estimated by multiplying the self-reported frequency and portion size of each food item with the corresponding nutritional content, obtained from data sources provided by the US Department of Agriculture. In the present study, we used AHEI-2010 to assess adherence to healthy dietary patterns (ranged from 0 to 100), based on previous literature (12-13). The mean of the last 2 blood pressure measurements was used. Fasting blood samples were also collected for further laboratory assays (e.g., plasma fasting glucose [PFG]). Total cholesterol and triglycerides were measured by enzymatic methods. High-density lipoprotein (HDL) cholesterol was assayed after dextran sulfate–magnesium precipitation, and LDL-C was estimated from the Friedewald equation (14).

| **Supplementary Table S1.** List of 43 genetic variants used in the LPA polygenetic risk score. | | | | | |
| --- | --- | --- | --- | --- | --- |
| **rsID** | **Chromosome: Position (GRCh37/hg19)** | **Effect allele** | **Other allele** | **Conditional association with Lp(a), nmol/l Beta^*^** | **Conditional association with Lp(a), nmol/l SE^*^** |
| rs74617384 | 6:160997118 | T | A | 91.160 | 1.075 |
| rs140570886 | 6:161013013 | C | T | 172.430 | 1.720 |
| rs73596816 | 6:161017363 | A | G | 41.280 | 1.290 |
| rs182443492 | 6:160891897 | A | C | 79.120 | 2.150 |
| rs369686024 | 6:161032800 | A | G | 41.280 | 1.720 |
| rs56393506 | 6:161089307 | T | C | 26.660 | 0.860 |
| rs151135411 | 6:160831796 | A | G | 149.425 | 5.805 |
| rs145099029 | 6:161292838 | C | A | 38.270 | 3.870 |
| rs41267813 | 6:160998199 | A | G | -126.420 | 6.235 |
| rs6916433 | 6:160890350 | T | A | -10.105 | 0.645 |
| rs783147 | 6:161137990 | A | G | -4.300 | 0.645 |
| rs41266379 | 6:160953137 | C | T | 15.265 | 1.505 |
| rs143461353 | 6:160954800 | T | C | 28.165 | 2.150 |
| rs142126734 | 6:160942926 | A | G | 16.125 | 1.075 |
| rs139609547^#^ | 6:160899049 | A | - | 9.460 | 0.860 |
| rs1835346 | 6:161162290 | G | A | 11.180 | 1.505 |
| rs4252152 | 6:161159366 | G | T | 19.565 | 1.935 |
| rs79246098 | 6:161078894 | C | T | 13.330 | 1.935 |
| rs139145675 | 6:160966559 | A | G | -48.375 | 5.160 |
| rs41259144 | 6:161022107 | T | C | -20.640 | 1.720 |
| rs9456551 | 6:161012805 | C | T | 7.740 | 0.430 |
| rs41267809 | 6:160953642 | G | A | -14.190 | 1.290 |
| rs34371670 | 6:161257953 | T | C | -18.060 | 1.505 |
| rs41269876 | 6:161070653 | A | C | -17.630 | 1.290 |
| rs141834709 | 6:160909667 | A | T | 18.705 | 2.150 |
| rs4252170 | 6:161162406 | C | T | 6.880 | 0.860 |
| rs138491411 | 6:161251940 | G | A | 10.750 | 1.720 |
| rs183815886 | 6:160720804 | C | G | 31.820 | 3.870 |
| rs117446263 | 6:160847571 | A | G | -11.180 | 1.290 |
| rs200684404 | 6:160543317 | T | C | 145.555 | 19.780 |
| rs200144324 | 6:160493099 | T | C | 175.225 | 24.295 |
| rs77337569 | 6:161087652 | G | T | 11.180 | 1.720 |
| rs186418835 | 6:161214526 | A | G | -20.855 | 3.225 |
| rs117534432 | 6:161177443 | T | C | 7.095 | 1.075 |
| rs200376184 | 6:161011999 | C | G | 37.625 | 5.805 |
| rs11753588 | 6:161189071 | A | G | -5.160 | 0.645 |
| rs4709474 | 6:161285760 | G | A | 3.655 | 0.430 |
| rs191690882 | 6:161031132 | A | G | -28.380 | 4.085 |
| rs182349273 | 6:161255668 | G | A | 73.960 | 12.255 |
| rs75274517 | 6:161088956 | A | G | -13.975 | 2.150 |
| rs143365644 | 6:160825930 | T | A | 7.955 | 1.075 |
| rs139389770 | 6:161135746 | G | T | -11.180 | 1.935 |
| rs140606700 | 6:161250301 | G | A | 13.760 | 2.580 |
| ^*^ Weights for the association between the 43 genetic variants and lipoprotein(a) were acquired from Burgess et al. (2018)^1^. ^#^ Effect allele for rs139609547 was obtained from dbSNP by https://www.ncbi.nlm.nih.gov/snp/?term=rs139609547. | | | | | |

**Supplementary Table S2.** Definitions of risk factor within target ranges in UK Biobank.

| **Risk factor within target ranges** | **Definition** |
| --- | --- |
| Ideal BMI | BMI ≥18.5 kg/m^2^ BUT <30 kg/m^2^ |
| Nonsmoking | Never smoking OR having quit smoking for at least 10 years before the recruitment for non-disease reasons. |
| Ideal diet quality | Healthy diet score was calculated by the following dietary factors:   1. Red meat <3.5 servings/week; 2. Processed meat <1 serving/week; 3. Fresh fruit ≥3 servings/day OR fresh vegetables ≥3 servings/day OR a combination ≥4.5 servings/day; 4. Whole grain ≥3 servings/day; 5. Refined grain <1.5 servings/day; 6. Fish ≥2 servings/week; 7. Never adding salt to food   **Each ideal dietary factor assigns one score, resulting in a healthy diet score ranging from 0 to 7. An ideal diet quality was defined as a healthy diet score of ≥4.** |
| Ideal sleep duration | Sleep duration ≥7 h/d BUT <9 h/d |
| Regular exercise | Engaging in at least 150 min/wk of moderate activity OR 75 min/wk of vigorous activity per week (or an equivalent combination), OR moderate activity at least 5 d/wk OR vigorous activity at least 3 d/wk (≥10 minutes continuously at a time) |
| Ideal glycemic control | HbA1c <48 mmol/mol (6.5%) |
| Ideal BP control | SBP <140 mmHg & DBP <90 mmHg |
| Ideal LDL-C control | LDL-C <4 mmol/L (15) |

Abbreviations: BMI, body mass index; BP, blood pressure; DBP, diastolic blood pressure; HbA1c, glycated hemoglobin; SBP, systolic blood pressure; LDL-C, low-density lipoprotein cholesterol.

**Supplementary Table S3.** Definitions of risk factor within target ranges in ARIC.

| **Risk factor within target ranges** | **Definition** |
| --- | --- |
| Ideal BMI | BMI ≥18.5 kg/m^2^ BUT <30 kg/m^2^ |
| Nonsmoking | Not current smokers |
| Ideal diet quality | Alternative Healthy Eating Index-2010 (AHEI-2010) was used to assess adherence to healthy dietary patterns. The AHEI-2010 score was created to include foods, beverages, and nutrients associated with chronic diseases, based on a previous literature (13).  **An ideal diet quality was defined as an AHEI-2010 score of ≥51.05 (median).** |
| Regular exercise | At least one physical activity reported for a total duration of at least 1 hour per week for 10 or more months per year (16). |
| Ideal glycemic control | FPG <7.0 mmol/l |
| Ideal BP control | SBP <140 mmHg & DBP <90 mmHg |
| Ideal LDL-C control | LDL-C <4 mmol/L (15) |

Abbreviations: BMI, body mass index; BP, blood pressure; DBP, systolic blood pressure; SBP, systolic blood pressure; LDL-C, low-density lipoprotein cholesterol.

**Supplementary Table S4.** ICD and OCPS codes used to define ASCVD in the UK Biobank.

| **ASCVD outcomes** | **ICD-9 Diagnosis Codes** | **ICD-10 Diagnosis Codes** | **OCPS-4 (operations)** |
| --- | --- | --- | --- |
| MI | 410 Acute myocardial infarction  411 Other acute and subacute forms of  ischaemic heart disease  412 Old myocardial infarction  42979 Ill-defined descriptions and complications of heart disease - Other | I21 Acute myocardial infarction  I22 Subsequent myocardial infarction  I23 Certain current complications following acute myocardial infarction  I24.1 Dressler syndrome  I25.2 Old myocardial infarction | N/A |
| PAD | 440 Atherosclerosis  444 Arterial embolism and thrombosis  4438 Other specified peripheral vascular disease  4439 Peripheral vascular disease, unspecified | I70 Atherosclerosis  I74 Arterial embolism and thrombosis  I73.8 Other specified peripheral vascular diseases  I73.9 Peripheral vascular disease, unspecified | L50 Other emergency bypass of iliac artery  L51 Other bypass of iliac artery  L52 Reconstruction of iliac artery  L54 Transluminal operations on iliac artery  L58 Other emergency bypass of femoral artery  L59 Other bypass of femoral artery  L60 Reconstruction of femoral artery  L63 Transluminal operations on femoral artery  X09 Amputation of leg |
| IS | 434 Occlusion of cerebral arteries  436 Acute, but ill-defined, cerebrovascular disease | I63 Cerebral infarction  I64 Stroke, not specified as haemorrhage or infarction. | N/A |
| CVD mortality | N/A | Any diagnosis code pertaining to mortality causes associated with Chapter IX, which encompasses diseases of the circulatory system | N/A |

Abbreviations: ASCVD, arteriosclerotic cardiovascular disease; CVD, cardiovascular disease; MI, myocardial infarction; PAD, peripheral arterial disease; IS, ischemic stroke. ICD, International Classification of Diseases; OCPS, Office of Population Censuses and Surveys Classification of Interventions and Procedures.

**Supplementary Table S5.** Characteristics of individuals with low Lp(a) PRS and individuals with high Lp(a) PRS in UK Biobank.

| **Characteristics** | **Control**  **(Quintile 1- 4)** | **Participants with high Lp(a) PRS (Quintile 5)** | | | | |
| --- | --- | --- | --- | --- | --- | --- |
|  |  | **Overall** | **Risk factor within target ranges** | | | |
|  |  |  | **0-2** | **3-4** | **5-6** | **7-8** |
| No. of participants | 219,245 (80.0) | 54,753 (20.0) | 807 (0.3) | 9,950 (3.6) | 27,615 (10.1) | 16,381 (6.0) |
| Lp(a) PRS* | -140.8 to 71.6 | 71.7 to 388.4 | 71.8 to 344.2 | 71.6 to 334.5 | 71.6 to 371.5 | 71.6 to 388.4 |
| Circulating Lp(a), nmol/l* | 3.8 to 189.0 | 3.8 to 189.0 | 4.0 to 188.6 | 3.8 to 189.0 | 3.8 to 189.0 | 3.8 to 189.0 |
| Age, y | 56.3±8.0 | 55.7±8.1 | 56.1±7.7 | 56.7±7.7 | 56.2±8.1 | 54.1±8.2 |
| Women | 125,597 (57.3) | 29,688 (54.2) | 336 (41.6) | 4,533 (45.6) | 14,513 (52.6) | 10,306 (62.9) |
| FHx of CVD | 89,291 (40.7) | 23,408 (42.8) | 363 (45.0) | 4,397 (44.2) | 11,861 (43.0) | 6,787 (41.4) |
| TDI | -1.6±2.9 | -1.6±2.9 | -0.6±3.2 | -1.2±3.1 | -1.6±2.9 | -1.9±2.7 |
| BMI, kg/m^2^ | 27.1±4.6 | 2.7±4.7 | 33.6±5.1 | 30.3±5.3 | 27.1±4.4 | 24.8±3.1 |
| Regular exercise | 168,702 (77.0) | 42,300 (77.3) | 165 (20.5) | 5,274 (53.0) | 21,475 (77.8) | 15,386 (93.9) |
| Healthy diet score | 3.7±1.4 | 3.7±1.4 | 2.4±1.0 | 2.9±1.2 | 3.6±1.4 | 4.4±1.2 |
| Non-smoking | 155,773 (71.1) | 39,068 (71.4) | 154 (19.1) | 4,729 (47.5) | 19,317 (70.0) | 14,868 (90.1) |
| Sleep duration, h/d | 7.2±1.1 | 7.2±1.0 | 6.7±1.6 | 7.0±1.4 | 7.2±1.0 | 7.3±0.7 |
| HbA1c, mmol/L | 35.5±5.8 | 35.5±6.0 | 42.3±12.6 | 37.6±8.4 | 35.3±5.3 | 34.1±3.9 |
| Antihyperglycemic drug use | 4,905 (2.2) | 1,288 (2.4) | 76 (9.4) | 525 (5.3) | 573 (2.1) | 114 (0.7) |
| SBP, mmHg | 139.4±19.5 | 139.1±19.5 | 152.2±15.9 | 148.3±18.1 | 141.0±19.4 | 129.4±16.2 |
| DBP, mmHg | 82.2±10.6 | 82.2±10.7 | 90.6±9.9 | 87.5±10.3 | 83.0±10.5 | 77.3±9.2 |
| Antihypertensive drug use | 36,316 (16.6) | 8,624 (15.8) | 230 (28.5) | 2,344 (23.6) | 4,554 (16.5) | 1,496 (9.1) |
| LDL-C, mmol/L | 3.6±0.8 | 3.6±0.8 | 4.2±0.9 | 4.0±0.9 | 3.7±0.8 | 3.3±0.6 |
| Statin use | 23,125 (10.6) | 5,403 (9.9) | 126 (15.6) | 1,254 (12.6) | 2,838 (10.3) | 1,185 (7.2) |
| hs-CRP, mg/L | 2.5±4.3 | 2.4±3.9 | 4.7±5.5 | 3.4±4.5 | 2.3±3.7 | 1.7±3.5 |
| WBC count, 10^9^/L | 6.8±2.0 | 6.8±2.0 | 8.0±3.3 | 7.3±1.9 | 6.8±1.8 | 6.4±2.0 |
| Platelet count, 10^9^/L | 254.7±58.6 | 252.1±58.2 | 261.9±60.9 | 256.5±60.6 | 252.5±58.2 | 248.3±56.1 |

Data are described as n (%) and mean±SD.

BMI, body mass index; CVD, cardiovascular disease; DBP, diastolic blood pressure; FHx, family history; HbA1c, glycosylated hemoglobin; hs-CRP, hypersensitive C-reactive protein; Lp(a), Lipoprotein(a); LDL-C, low density lipoprotein cholesterol; PRS, polygenetic risk score; SBP, systolic blood pressure; TDI, Townsend deprivation index; WBC, white blood cell

*The range for Lp(a) PRS and circulating Lp(a) concentration is provided. The identical range of circulating Lp(a) levels between the control group and the highest quintile of Lp(a) PRS is likely due to the fact that the Lp(a) PRS does not fully capture the true circulating Lp(a) levels. Certain medications or pathological conditions may also influence circulating Lp(a) levels, leading to discrepancies between genetic risk (PRS) and measured Lp(a) levels.

**Supplementary Table S6.** Characteristics of individuals with non-elevated circulating Lp(a) and individuals with elevated circulating Lp(a) in ARIC.

| **Characteristics** | **Control**  **(Quintile 1- 4)** | **Participants with elevated circulating Lp(a) (Quintile 5)** | | | |
| --- | --- | --- | --- | --- | --- |
|  |  | **Overall** | **Risk factor within target ranges** | | |
|  |  |  | **0-3** | **4-5** | **6-7** |
| Participants | 9,234 | 2,290 | 415 | 1,227 | 648 |
| Circulating Lp(a), mg/dL* | 0.3 to 71.4 | 40.5 to 245.1 | 40.5 to 209.1 | 40.5 to 245.1 | 40.5 to 200.1 |
| Age, y | 53.9±5.7 | 53.9±5.7 | 54.8±5.9 | 54.0±5.7 | 53.7±5.5 |
| Women | 4,921 (53.3) | 1,397 (61.0) | 247 (59.5) | 735 (59.9) | 415 (64.0) |
| FHx of CVD | 2,991 (32.4) | 841 (36.7) | 161 (38.8) | 453 (36.9) | 227 (35.0) |
| Family income, ≥$50,000 | 2,476 (26.8) | 602 (26.3) | 51 (12.3) | 309 (25.2) | 242 (37.4) |
| Ethnicity |  |  |  |  |  |
| White | 7,042 (76.3) | 1,743 (76.1) | 240 (57.8) | 932 (76.0) | 571 (88.1) |
| Black | 2,192 (23.7) | 547 (23.9) | 175 (42.2) | 295 (24.0) | 77 (11.9) |
| BMI, kg/m^2^ | 27.5±5.2 | 27.5±5.5 | 30.9±6.4 | 27.5±5.4 | 25.5±3.6 |
| Regular exercise | 2,960 (32.1) | 742 (32.4) | 21 (5.1) | 273 (22.3) | 448 (69.1) |
| AHEI-2010 | 62.6±8.8 | 63.5±9.1 | 59.0±8.2 | 62.4±8.8 | 68.5±8.0 |
| Non-smoking | 6,859 (74.3) | 1,754 (76.6) | 217 (52.3) | 924 (75.3) | 613 (94.6) |
| PFG, mmol/L | 107.0±35.7 | 106.0±35.7 | 127.7±63.5 | 103.2±26.1 | 97.6±15.6 |
| Antihyperglycemic drug use | 390 (4.2) | 98 (4.3) | 50 (12.1) | 43 (3.5) | 5 (0.8) |
| SBP, mmHg | 120.7±18.4 | 120.3±18.7 | 132.9±22.3 | 119.6±17.6 | 113.6±13.7 |
| DBP, mmHg | 73.7±11.1 | 73.3±11.4 | 78.7±13.4 | 73.1±11.0 | 70.0±9.2 |
| Antihypertensive drug use | 2,020 (21.9) | 518 (22.6) | 159 (38.3) | 279 (22.7) | 80 (12.4) |
| LDL-C, mmol/L | 3.5±1.0 | 3.8±1.0 | 4.4±1.0 | 3.9±1.0 | 3.4±0.7 |
| Statin use | 31 (0.3) | 15 (0.7) | 3 (0.7) | 9 (0.7) | 3 (0.5) |
| WBC count, 10^9^/L | 6.1±1.9 | 6.0±2.1 | 6.7±2.2 | 6.1±2.2 | 5.6±1.7 |
| Platelet count, 10^9^/L | 257.1±64.8 | 260.7±64.1 | 263.9±64.4 | 261.6±62.9 | 258.9±65.2 |

Data are described as n (%) and mean±SD.

Abbreviations: AHEI, alternative Healthy Eating Index; BMI, body mass index; CVD, cardiovascular disease; FHx, family history; HbA1c, glycosylated hemoglobin; Lp(a), Lipoprotein(a); LDL-C, low density lipoprotein cholesterol; PFG, plasma fasting glucose; SBP, systolic blood pressure; DBP, diastolic blood pressure; WBC, white blood cell.

*The range for circulating Lp(a) is provided. The quintiles of circulating Lp(a) levels were determined using ethnicity-specific cutoffs to account for variations in Lp(a) distributions across different ethnic groups. As a result, some overlap in Lp(a) levels between quintile 1- quintile 4 and quintile 5 may occur.

**Supplementary Table S7.** Associations of circulating Lp(a) or Lp(a) PRS with risk of ASCVD in UK Biobank and ARIC, according to quintiles.

|  | **Circulating Lp(a) in UK Biobank** | | **Lp(a) PRS in UK Biobank** | | **Circulating Lp(a) in ARIC** | |
| --- | --- | --- | --- | --- | --- | --- |
|  | **Events/person-y** | **HR (95% CI)** | **Events/person-y** | **HR (95% CI)** | **Events/person-y** | **HR (95% CI)** |
| **ASCVD** |  |  |  |  |  |  |
| Lp(a) Q1-Q4 | 12,592/2,829,517 | Reference | 12,090/2,683,157 | Reference | 2,457/206,282 | Reference |
| Lp(a) Q5 | 3,725/704,658 | 1.21 (1.17, 1.26) | 3,504/668,472 | 1.18 (1.13, 1.22) | 700/49,738 | 1.22 (1.12, 1.32) |
| **MI** |  |  |  |  |  |  |
| Lp(a) Q1-Q4 | 5,564/2,868,812 | Reference | 5,321/2,720,392 | Reference | 1,238/211,485 | Reference |
| Lp(a) Q5 | 1,826/715,102 | 1.32 (1.25, 1.39) | 1,692/678,443 | 1.26 (1.20, 1.33) | 375/51,225 | 1.28 (1.14, 1.44) |
| **PAD** |  |  |  |  |  |  |
| Lp(a) Q1-Q4 | 2,173/2,885,466 | Reference | 2,083/2,720,392 | Reference | 344/233,047 | Reference |
| Lp(a) Q5 | 673/720,899 | 1.27 (1.17, 1.39) | 638/678,443 | 1.25 (1.14, 1.36) | 104/57,121 | 1.29 (1.03, 1.61) |
| **IS** |  |  |  |  |  |  |
| Lp(a) Q1-Q4 | 3,135/2,881,759 | Reference | 3,037/2,720,392 | Reference | 695/215,670 | Reference |
| Lp(a) Q5 | 835/720,082 | 1.09 (1.01, 1.18) | 772/678,443 | 1.04 (0.96, 1.13) | 208/52,606 | 1.22 (1.05, 1.43) |
| **CVD death** |  |  |  |  |  |  |
| Lp(a) Q1-Q4 | 4,055/2,873,238 | Reference | 3,915/2,725,201 | Reference | 1,070/221,983 | Reference |
| Lp(a) Q5 | 1,107/71,8427 | 1.13 (1.05, 1.20) | 1,044/681,294 | 1.08 (1.02, 1.16) | 291/54,304 | 1.13 (1.01, 1.29) |

For circulating Lp(a) in UK Biobank, the model was adjusted for age, sex, ethnicity, TDI, FHx of CVD, BMI, regular exercise, healthy diet score, smoking, sleep duration, HbA1c, SBP, DBP, LDL-C, antihyperglycemic drug use, antihypertensive drug use, and statin use.

For Lp(a) PRS in UK Biobank, the analyses were conducted among the White only. The model was adjusted for the covariates that were used for circulating Lp(a) and further adjusted for the first 10 principal components of ancestry.

For circulating Lp(a) in ARIC, the model was adjusted for age, sex, ethnicity, family income, FHx of CVD, BMI, regular exercise, AHEI-2010, smoking, FPG, SBP, DBP, LDL-C, antihyperglycemic drug use, antihypertensive drug use, and statin use.

Abbreviations: ASCVD, arteriosclerotic cardiovascular disease; ARIC, Atherosclerosis Risk In Communities; CVD, cardiovascular disease; CI, confidence interval; HR, hazard ratio; MI, myocardial infarction; PAD, peripheral arterial disease; IS, ischemic stroke.

**Supplementary Table S8.** Associations of individual risk factor within target range with risk of ASCVD among participants with elevated circulating Lp(a) or high Lp(a) PRS in UK Biobank.

|  | **Participants with elevated circulating Lp(a)** | | **Participants with high Lp(a) PRS** | |
| --- | --- | --- | --- | --- |
| **ASCVD** | **Events/person-y** | **HR (95% CI)** | **Events/person-y** | **HR (95% CI)** |
| Ideal BMI | 2,576/545,953 | 0.78 (0.72, 0.83) | 2,394/516,590 | 0.80 (0.74, 0.86) |
| Regular exercise | 2,769/545,872 | 0.89 (0.83, 0.96) | 2,591/516,999 | 0.89 (0.82, 0.96) |
| Ideal diet quality | 1,774/395,681 | 0.86 (0.80, 0.91) | 1,626/367,422 | 0.87 (0.82, 0.94) |
| Non-smoking | 2,294/506,793 | 0.64 (0.60, 0.69) | 2,149/480,017 | 0.65 (0.60, 0.69) |
| Ideal sleep duration | 2,406/490,339 | 0.87 (0.82, 0.93) | 2,238/468,446 | 0.84 (0.78, 0.90) |
| Ideal glycemic control | 3,454/686,800 | 0.67 (0.56, 0.80) | 3,249/651,298 | 0.76 (0.63, 0.90) |
| Ideal BP control | 1,290/361,631 | 0.78 (0.73, 0.84) | 1,184/344,370 | 0.76 (0.71, 0.82) |
| Ideal LDL-C control | 2,335/469,605 | 0.77 (0.72, 0.83) | 2,284/464,496 | 0.76 (0.70, 0.82) |
| **MI** | **Events/person-y** | **HR (95% CI)** | **Events/person-y** | **HR (95% CI)** |
| Ideal BMI | 1,268/553591 | 0.81 (0.77, 0.86) | 1,175/523,792 | 0.83 (0.75, 0.93) |
| Regular exercise | 1,371/553760 | 0.90 (0.85, 0.94) | 1,260/524,567 | 0.90 (0.81, 1.01) |
| Ideal diet quality | 873/401072 | 0.83 (0.79, 0.87) | 777/372,512 | 0.88 (0.80, 0.97) |
| Non-smoking | 1,153/513666 | 0.71 (0.68, 0.75) | 1,060/486,492 | 0.69 (0.62, 0.76) |
| Ideal sleep duration | 1,176/497290 | 0.85 (0.81, 0.89) | 1,073/475,099 | 0.81 (0.73, 0.90) |
| Ideal glycemic control | 1,698/696686 | 0.70 (0.62, 0.80) | 1,584/660,728 | 0.75 (0.57, 0.98) |
| Ideal BP control | 631/366129 | 0.79 (0.75, 0.83) | 568/348,651 | 0.72 (0.65, 0.80) |
| Ideal LDL-C control | 1,057/476470 | 0.67 (0.63, 0.70) | 1,009/471,320 | 0.62 (0.56, 0.69) |
| **PAD** | **Events/person-y** | **HR (95% CI)** | **Events/person-y** | **HR (95% CI)** |
| Ideal BMI | 450/557,782 | 0.89 (0.82, 0.96) | 423/527,575 | 0.83 (0.70, 0.99) |
| Regular exercise | 498/558,277 | 0.81 (0.75, 0.88) | 463/528,585 | 0.87 (0.73, 1.04) |
| Ideal diet quality | 311/403,899 | 0.82 (0.76, 0.89) | 287/374,966 | 0.84 (0.72, 0.99) |
| Non-smoking | 341/517,643 | 0.42 (0.39, 0.45) | 327/490,153 | 0.44 (0.38, 0.52) |
| Ideal sleep duration | 436/501,143 | 0.81 (0.75, 0.87) | 411/478,519 | 0.92 (0.78, 1.08) |
| Ideal glycemic control | 602/702,281 | 0.62 (0.52, 0.74) | 566/665,822 | 0.67 (0.47, 0.96) |
| Ideal BP control | 227/368,098 | 0.88 (0.81, 0.95) | 223/350,330 | 0.85 (0.72, 1.01) |
| Ideal LDL-C control | 446/479,409 | 0.97 (0.89, 1.06) | 446/474,003 | 0.83 (0.69, 0.99) |
| **IS** | **Events/person-y** | **HR (95% CI)** | **Events/person-y** | **HR (95% CI)** |
| Ideal BMI | 605/557,035 | 0.87 (0.81, 0.94) | 555/526,860 | 0.90 (0.76, 1.06) |
| Regular exercise | 623/557,581 | 0.91 (0.85, 0.98) | 575/527,821 | 0.87 (0.74, 1.02) |
| Ideal diet quality | 404/403,530 | 0.90 (0.85, 0.96) | 390/374,404 | 0.98 (0.87, 1.13) |
| Non-smoking | 546/516,632 | 0.76 (0.72, 0.82) | 499/489,141 | 0.74 (0.63, 0.85) |
| Ideal sleep duration | 553/500,537 | 0.91 (0.85, 0.97) | 502/477,918 | 0.87 (0.75, 1.01) |
| Ideal glycemic control | 777/701,376 | 0.75 (0.63, 0.89) | 720/664,907 | 0.81 (0.54, 1.20) |
| Ideal BP control | 263/367,894 | 0.72 (0.67, 0.77) | 247/350,086 | 0.68 (0.58, 0.80) |
| Ideal LDL-C control | 540/479,058 | 0.97 (0.90, 1.04) | 524/473,625 | 0.91 (0.77, 1.06) |
| **CVD death** | **Events/person-y** | **HR (95% CI)** | **Events/person-y** | **HR (95% CI)** |
| Ideal BMI | 720/555,595 | 0.71 (0.67, 0.75) | 668/525,424 | 0.70 (0.61, 0.80) |
| Regular exercise | 777/556,323 | 0.80 (0.75, 0.85) | 757/526,610 | 0.85 (0.74, 0.97) |
| Ideal diet quality | 523/402,231 | 0.86 (0.81, 0.91) | 475/373,362 | 0.88 (0.77, 0.99) |
| Non-smoking | 637/515,250 | 0.58 (0.54, 0.61) | 622/487,788 | 0.62 (0.55, 0.70) |
| Ideal sleep duration | 708/499,297 | 0.84 (0.79, 0.89) | 673/476,674 | 0.90 (0.80, 1.03) |
| Ideal glycemic control | 997/699,620 | 0.64 (0.56, 0.73) | 940/663,258 | 0.64 (0.48, 0.86) |
| Ideal BP control | 388/366,164 | 0.84 (0.79, 0.89) | 348/348,496 | 0.86 (0.75, 0.98) |
| Ideal LDL-C control | 758/477,791 | 0.98 (0.92, 1.05) | 735/472,370 | 0.91 (0.79, 1.05) |

For participants with elevated circulating Lp(a), the model was adjusted for Lp(a), age, sex, ethnicity, TDI, FHx of CVD, antihyperglycemic drug use, antihypertensive drug use, statin use and the remaining individual risk factors.

For participants with high Lp(a) PRS, the analyses were conducted among the White only. The model was adjusted for the covariates that were used for circulating Lp(a) and further adjusted for the first 10 principal components of ancestry.

Abbreviations: ASCVD, arteriosclerotic cardiovascular disease; BMI, body mass index; BP, blood pressure; CVD, cardiovascular disease; CI, confidence interval; HR, hazard ratio; LDL-C, low-density lipoprotein cholesterol; Lp(a), lipoprotein(a); MI, myocardial infarction; PAD, peripheral arterial disease; PRS, polygenetic risk score; IS, ischemic stroke.

**Supplementary Table S9.** Associations of individual risk factor within target range with risk of ASCVD among participants with elevated circulating Lp(a) in ARIC.

| **ASCVD** | **Events/person-y** | **HR (95% CI)** |
| --- | --- | --- |
| Ideal BMI | 457/36,727 | 0.86 (0.73, 1.01) |
| Regular exercise | 192/16,906 | 0.86 (0.80, 1.11) |
| Ideal diet quality | 342/26,972 | 0.93 (0.80, 1.09) |
| Non-smoking | 497/39,948 | 0.51 (0.43, 0.61) |
| Ideal glycemic control | 596/47,027 | 0.53 (0.40, 0.70) |
| Ideal BP control | 518/42,925 | 0.61 (0.51, 0.73) |
| Ideal LDL-C control | 364/31,165 | 0.75 (0.64, 0.87) |
| **MI** | **Events/person-y** | **HR (95% CI)** |
| Ideal BMI | 252/37,537 | 0.93 (0.73, 1.17) |
| Regular exercise | 98/17,273 | 0.81 (0.64, 1.04) |
| Ideal diet quality | 181/27,338 | 0.95 (0.77, 1.17) |
| Non-smoking | 266/40,545 | 0.58 (0.45, 0.73) |
| Ideal glycemic control | 316/48,053 | 0.58 (0.40, 0.85) |
| Ideal BP control | 274/43,764 | 0.57 (0.44, 0.72) |
| Ideal LDL-C control | 179/31,888 | 0.63 (0.51, 0.76) |
| **PAD** | **Events/person-y** | **HR (95% CI)** |
| Ideal BMI | 69/42,134 | 1.04 (0.67, 1.63) |
| Regular exercise | 27/19,149 | 1.11 (0.70, 1.86) |
| Ideal diet quality | 51/30,516 | 0.93 (0.62, 1.38) |
| Non-smoking | 49/45,531 | 0.17 (0.11, 0.25) |
| Ideal glycemic control | 82/53,791 | 0.69 (0.34, 1.37) |
| Ideal BP control | 72/48,981 | 0.54 (0.34, 0.85) |
| Ideal LDL-C control | 56/35,305 | 0.91 (0.61, 1.35) |
| **IS** | **Events/person-y** | **HR (95% CI)** |
| Ideal BMI | 129/38,659 | 0.81 (0.60, 1.12) |
| Regular exercise | 51/17,713 | 0.79 (0.57, 1.10) |
| Ideal diet quality | 106/28,011 | 1.02 (0.77, 1.36) |
| Non-smoking | 152/41,576 | 0.63 (0.45, 0.84) |
| Ideal glycemic control | 172/49,304 | 0.43 (0.27, 0.68) |
| Ideal BP control | 152/44,966 | 0.66 (0.47, 0.90) |
| Ideal LDL-C control | 112/32,356 | 0.83 (0.63, 1.09) |
| **CVD death** | **Events/person-y** | **HR (95% CI)** |
| Ideal BMI | 178/39,784 | 0.71 (0.55, 0.92) |
| Regular exercise | 79/18,134 | 0.88 (0.69, 1.15) |
| Ideal diet quality | 138/28,754 | 0.88 (0.70, 1.13) |
| Non-smoking | 208/42,762 | 0.48 (0.36, 0.62) |
| Ideal glycemic control | 236/50,771 | 0.41 (0.27, 0.62) |
| Ideal BP control | 202/46,224 | 0.48 (0.37, 0.62) |
| Ideal LDL-C control | 145/33,297 | 0.75 (0.59, 0.94) |

The model was adjusted for age, sex, ethnicity, household income, FHx of CVD, antihyperglycemic drug use, antihypertensive drug use, statin use, and the remaining risk factors.

Abbreviations: ASCVD, arteriosclerotic cardiovascular disease; ARIC, Atherosclerosis Risk In Communities; BMI, body mass index; BP, blood pressure; CVD, cardiovascular disease; CI, confidence interval; HR, hazard ratio; LDL-C, low density lipoprotein cholesterol; Lp(a), lipoprotein(a); MI, myocardial infarction; PAD, peripheral arterial disease; IS, ischemic stroke.

**Supplementary Table S10.** Associations of number of risk factor within target ranges with risk of individual ASCVD outcomes among participants with elevated circulating Lp(a) or high Lp(a) PRS in UK Biobank.

|  | **Participants with elevated circulating Lp(a)** | | **Participants with high Lp(a) PRS** | |
| --- | --- | --- | --- | --- |
| **ASCVD** | **Events/person-y** | **HR (95% CI)** | **Events/person-y** | **HR (95% CI)** |
| 0-2 risk factor within target ranges | 126/9807 | Reference | 122/9354 | Reference |
| 3-4 risk factor within target ranges | 1,120/125808 | 0.72 (0.60, 0.86) | 1,068/118491 | 0.71 (0.59, 0.86) |
| 5-6 risk factor within target ranges | 1,911/360235 | 0.48 (0.40, 0.57) | 1,781/336801 | 0.47 (0.39, 0.57) |
| 7-8 risk factor within target ranges | 568/208806 | 0.30 (0.25, 0.37) | 422/203824 | 0.30 (0.24, 0.36) |
| Per 1 target increment | - | 0.79 (0.78, 0.81) | - | 0.79 (0.77, 0.81) |
| *P* for linear trend | - | <0.001 | - | <0.001 |
| **MI** | **Events/person-y** | **HR (95% CI)** | **Events/person-y** | **HR (95% CI)** |
| 0-2 risk factor within target ranges | 63/10061 | Reference | 62/9595 | Reference |
| 3-4 risk factor within target ranges | 552/128337 | 0.72 (0.56, 0.94) | 524/120900 | 0.70 (0.54, 0.91) |
| 5-6 risk factor within target ranges | 951/365573 | 0.49 (0.38, 0.63) | 865/341863 | 0.46 (0.35, 0.59) |
| 7-8 risk factor within target ranges | 260/211128 | 0.28 (0.21, 0.37) | 241/206083 | 0.26 (0.20, 0.35) |
| Per 1 target increment | - | 0.78 (0.76, 0.81) | - | 0.78 (0.75, 0.80) |
| *P* for linear trend | - | <0.001 | - | <0.001 |
| **PAD** | **Events/person-y** | **HR (95% CI)** | **Events/person-y** | **HR (95% CI)** |
| 0-2 risk factor within target ranges | 29/10240 | Reference | 24/9775 | Reference |
| 3-4 risk factor within target ranges | 232/129895 | 0.69 (0.47, 1.02) | 229/122361 | 0.83 (0.54, 1.26) |
| 5-6 risk factor within target ranges | 316/368752 | 0.38 (0.26, 0.56) | 289/344699 | 0.45 (0.29, 0.68) |
| 7-8 risk factor within target ranges | 96/212011 | 0.26 (0.17, 0.40) | 96/206856 | 0.33 (0.21, 0.52) |
| Per 1 target increment | - | 0.76 (0.72, 0.80) | - | 0.77 (0.73, 0.81) |
| *P* for linear trend | - | <0.001 | - | <0.001 |
| **IS** | **Events/person-y** | **HR (95% CI)** | **Events/person-y** | **HR (95% CI)** |
| 0-2 risk factor within target ranges | 25/10258 | Reference | 21/9800 | Reference |
| 3-4 risk factor within target ranges | 229/129845 | 0.75 (0.49, 1.13) | 209/122363 | 0.81 (0.51, 1.27) |
| 5-6 risk factor within target ranges | 442/368158 | 0.56 (0.37, 0.84) | 407/344049 | 0.62 (0.40, 0.97) |
| 7-8 risk factor within target ranges | 139/211819 | 0.38 (0.25, 0.58) | 135/206640 | 0.44 (0.28, 0.70) |
| Per 1 target increment | - | 0.83 (0.79, 0.87) | - | 0.85 (0.81, 0.89) |
| *P* for linear trend | - | <0.001 | - | <0.001 |
| **CVD death** | **Events/person-y** | **HR (95% CI)** | **Events/person-y** | **HR (95% CI)** |
| 0-2 risk factor within target ranges | 43/10305 | Reference | 37/9822 | Reference |
| 3-4 risk factor within target ranges | 369/129864 | 0.74 (0.54, 1.02) | 339/122403 | 0.79 (0.56, 1.11) |
| 5-6 risk factor within target ranges | 530/367441 | 0.43 (0.32, 0.59) | 511/343424 | 0.51 (0.36, 0.71) |
| 7-8 risk factor within target ranges | 165/210815 | 0.31 (0.22, 0.44) | 157/205644 | 0.36 (0.25, 0.51) |
| Per 1 target increment | - | 0.78 (0.75, 0.82) | - | 0.80 (0.76, 0.84) |
| *P* for linear trend | - | <0.001 | - | <0.001 |

For participants with elevated circulating Lp(a) in UK Biobank, the model was adjusted for Lp(a), age, sex, ethnicity, TDI, FHx of CVD, antihyperglycemic drug use, antihypertensive drug use, statin use and the remaining individual risk factors.

For participants with high Lp(a) PRS in UK Biobank, the analyses were conducted among the White only. The model was adjusted for the covariates that were used for circulating Lp(a) and further adjusted for the first 10 principal components of ancestry.

Abbreviations: ASCVD, arteriosclerotic cardiovascular disease; CVD, cardiovascular disease; CI, confidence interval; HR, hazard ratio; Lp(a), lipoprotein(a); MI, myocardial infarction; PAD, peripheral arterial disease; PRS, polygenetic risk score; IS, ischemic stroke.

**Supplementary Table S11.** Associations of number of risk factor within target ranges with risk of ASCVD among participants with elevated circulating Lp(a) in ARIC.

| **ASCVD** | **Events/person-y** | **HR (95% CI)** |
| --- | --- | --- |
| 0-3 risk factor within target ranges | 204/7,408 | Reference |
| 4-5 risk factor within target ranges | 374/26,405 | 0.60 (0.50, 0.72) |
| 6-7 risk factor within target ranges | 122/15,893 | 0.37 (0.29, 0.46) |
| Per 1 target increment | - | 0.74 (0.70, 0.80) |
| *P* for linear trend | - | <0.001 |
| **MI** | **Events/person-y** | **HR (95% CI)** |
| 0-3 risk factor within target ranges | 109/7,408 | Reference |
| 4-5 risk factor within target ranges | 212/26,435 | 0.67 (0.53, 0.86) |
| 6-7 risk factor within target ranges | 54/15,893 | 0.32 (0.23, 0.46) |
| Per 1 target increment | - | 0.73 (0.67, 0.79) |
| *P* for linear trend | - | <0.001 |
| **PAD** | **Events/person-y** | **HR (95% CI)** |
| 0-3 risk factor within target ranges | 40/8,929 | Reference |
| 4-5 risk factor within target ranges | 52/30,547 | 0.56 (0.36, 0.87) |
| 6-7 risk factor within target ranges | 12/17,644 | 0.27 (0.13, 0.53) |
| Per 1 target increment | - | 0.67 (0.57, 0.79) |
| *P* for linear trend | - | <0.001 |
| **IS** | **Events/person-y** | **HR (95% CI)** |
| 0-3 risk factor within target ranges | 68/8,143 | Reference |
| 4-5 risk factor within target ranges | 97/28,130 | 0.51 (0.37, 0.71) |
| 6-7 risk factor within target ranges | 43/16,332 | 0.46 (0.31, 0.70) |
| Per 1 target increment | - | 0.77 (0.69, 0.86) |
| *P* for linear trend | - | <0.001 |
| **CVD death** | **Events/person-y** | **HR (95% CI)** |
| 0-3 risk factor within target ranges | 92/8,671 | Reference |
| 4-5 risk factor within target ranges | 157/28,971 | 0.61 (0.46, 0.79) |
| 6-7 risk factor within target ranges | 42/16,661 | 0.31 (0.21, 0.46) |
| Per 1 target increment | - | 0.67 (0.62, 0.75) |
| *P* for linear trend | - | <0.001 |

The model was adjusted for Lp(a), age, sex, ethnicity, family income, FHx of CVD, antihyperglycemic drug use, antihypertensive drug use, and statin use.

Abbreviations: ASCVD, arteriosclerotic cardiovascular disease; ARIC, Atherosclerosis Risk In Communities; CVD, cardiovascular disease; CI, confidence interval; HR, hazard ratio; Lp(a), lipoprotein(a); MI, myocardial infarction; PAD, peripheral arterial disease; IS, ischemic stroke.

**Supplementary Table S12.** Incidence rates (per 1,000 person-y) for ASCVD according to the number of risk factor within target ranges among participants with elevated circulating Lp(a) or high Lp(a) PRS compared with those with non-elevated circulating Lp(a) or low Lp(a) PRS in UK Biobank and ARIC.

|  | **Events/ person-y** | **Incidence rate (95% CI)** | **Absolute rate difference (95% CI)** |
| --- | --- | --- | --- |
| **Circulating Lp(a) in UK Biobank** |  |  |  |
| 0-2 risk factor within target ranges | 126/9,807 | 12.85 (10.77, 14.93) | 8.40 (6.36, 10.47) |
| 3-4 risk factor within target ranges | 1,120/125,808 | 8.90 (8.45, 9.40) | 4.45 (4.04, 5.01) |
| 5-6 risk factor within target ranges | 1,911/360,235 | 5.30 (5.11, 5.53) | 0.85 (0.61, 1.09) |
| 7-8 risk factor within target ranges | 568/208,806 | 2.72 (2.50, 2.89) | -1.73 (-1.99, -1.54) |
| Control [Q1-Q4 Lp(a)] | 12,592/2,829,517 | 4.45 (4.37, 4.53) | Reference |
| **Lp(a) PRS in UK Biobank** |  |  |  |
| 0-2 risk factor within target ranges | 122/9,354 | 13.04 (10.70, 15.12) | 8.54 (6.17, 10.63) |
| 3-4 risk factor within target ranges | 1,068/118,491 | 9.01 (8.45, 9.56) | 4.51 (3.93, 5.06) |
| 5-6 risk factor within target ranges | 1,781/336,801 | 5.29 (5.10, 5.53) | 0.78 (0.60, 1.04) |
| 7-8 risk factor within target ranges | 422/203,824 | 2.61 (2.42, 2.77) | -1.89 (-2.08, -1.74) |
| Control [Q1-Q4 Lp(a)] | 12,090/2,683,157 | 4.51 (4.44, 4.56) | Reference |
| **Circulating Lp(a) in ARIC** |  |  |  |
| 0-3 risk factor within target ranges | 204/7,408 | 27.50 (24.84, 32.47) | 15.62 (12.72, 19.65) |
| 4-5 risk factor within target ranges | 374/26,405 | 14.15 (12.90, 15.79) | 2.24 (0.50, 3.81) |
| 6-7 risk factor within target ranges | 122/15,893 | 7.68 (6.45, 8.03) | -4.23 (-5.55, -2.84) |
| Control [Q1-Q4 Lp(a)] | 2,457/206,189 | 11.90 (11.60, 12.30) | Reference |

ARIC, Atherosclerosis Risk in Communities; CI, confidence interval; HR, hazard ratio; Lp(a), lipoprotein(a); PRS, polygenetic risk score

**Supplementary Table S13.** Incidence rates (per 1,000 person-y) for individual ASCVD outcomes according to the number of risk factor within target ranges among participants with elevated circulating Lp(a) compared with those with non-elevated circulating Lp(a) in UK Biobank.

|  | **Events/ person-y** | **Incidence rate (95% CI)** | **Absolute rate difference (95% CI)** |
| --- | --- | --- | --- |
| **MI** |  |  |  |
| 0-2 risk factor within target ranges | 63/10061 | 6.26 (4.63, 7.92) | 4.32 (2.72, 5.98) |
| 3-4 risk factor within target ranges | 552/128337 | 4.30 (4.00, 4.63) | 2.36 (2.05, 2.71) |
| 5-6 risk factor within target ranges | 951/365573 | 2.60 (2.46, 2.76) | 0.66 (0.51, 0.81) |
| 7-8 risk factor within target ranges | 260/211128 | 1.23 (1.10, 1.34) | -0.71 (-0.88, -0.58) |
| Control [Q1-Q4 circulating Lp(a)] | 5,564/2868812 | 1.94 (1.90, 1.99) | Reference |
| **PAD** |  |  |  |
| 0-2 risk factor within target ranges | 29/10240 | 2.83 (1.99, 3.84) | 2.08 (1.23, 3.10) |
| 3-4 risk factor within target ranges | 232/129895 | 1.79 (1.57, 2.03) | 1.03 (0.81, 1.28) |
| 5-6 risk factor within target ranges | 316/368752 | 0.86 (0.78, 0.95) | 0.10 (0.02, 0.20) |
| 7-8 risk factor within target ranges | 96/212011 | 0.45 (0.37, 0.54) | -0.30 (-0.38, -0.21) |
| Control [Q1-Q4 circulating Lp(a)] | 2,173/2885466 | 0.75 (0.72, 0.78) | Reference |
| **IS** |  |  |  |
| 0-2 risk factor within target ranges | 25/10258 | 2.44 (1.48, 3.36) | 1.35 (0.39, 2.29) |
| 3-4 risk factor within target ranges | 229/129845 | 1.76 (1.60, 1.97) | 0.68 (0.52, 0.90) |
| 5-6 risk factor within target ranges | 442/368158 | 1.20 (1.12, 1.32) | 0.11 (0.02, 0.24) |
| 7-8 risk factor within target ranges | 139/211819 | 0.66 (0.56, 0.77) | -0.43 (-0.53, -0.32) |
| Control [Q1-Q4 circulating Lp(a)] | 3,135/2881759 | 1.09 (1.05, 1.12) | Reference |
| **CVD death** |  |  |  |
| 0-2 risk factor within target ranges | 43/10305 | 4.17 (3.21, 5.23) | 2.76 (1.80, 3.81) |
| 3-4 risk factor within target ranges | 369/129864 | 2.84 (2.59, 3.12) | 1.43 (1.19, 1.73) |
| 5-6 risk factor within target ranges | 530/367441 | 1.44 (1.34, 1.58) | 0.03 (-0.07, 0.17) |
| 7-8 risk factor within target ranges | 165/210815 | 0.78 (0.67, 0.89) | -0.63 (-0.75, -0.51) |
| Control [Q1-Q4 circulating Lp(a)] | 4,055/2873238 | 1.41 (1.37, 1.46) | Reference |

Abbreviations: ASCVD, arteriosclerotic cardiovascular disease; CVD, cardiovascular disease; CI, confidence interval; Lp(a), lipoprotein(a); MI, myocardial infarction; PAD, peripheral arterial disease; IS, ischemic stroke.

**Supplementary Table S14.** Incidence rates (per 1,000 person-y) for ASCVD according to the number of risk factor within target ranges among participants with high Lp(a) PRS compared with those with low Lp(a) PRS in UK Biobank.

|  | **Events/person-y** | **Incidence rate (95% CI)** | **Absolute rate difference (95% CI)** |
| --- | --- | --- | --- |
| **MI** |  |  |  |
| 0-2 risk factor within target ranges | 62/9,595 | 6.46 (4.64, 7.77) | 4.51 (2.66, 5.85) |
| 3-4 risk factor within target ranges | 524/120,900 | 4.33 (3.92, 4.68) | 2.38 (1.97, 2.72) |
| 5-6 risk factor within target ranges | 865/341,863 | 2.53 (2.36, 2.71) | 0.57 (0.38, 0.74) |
| 7-8 risk factor within target ranges | 241/206,083 | 1.17 (1.05, 1.30) | -0.79 (-0.93, -0.64) |
| Control [Q1-Q4 Lp(a) PRS] | 5,321/2,720,392 | 1.96 (1.91, 2.00) | Reference |
| **PAD** |  |  |  |
| 0-2 risk factor within target ranges | 24/9,775 | 2.46 (1.44, 3.44) | 1.69 (0.69, 2.67) |
| 3-4 risk factor within target ranges | 229/122,361 | 1.87 (1.59, 2.12) | 1.11 (0.83, 1.35) |
| 5-6 risk factor within target ranges | 289/344,699 | 0.84 (0.76, 0.95) | 0.08 (-0.01, 0.20) |
| 7-8 risk factor within target ranges | 96/206,856 | 0.46 (0.37, 0.56) | -0.30 (-0.37, -0.20) |
| Control [Q1-Q4 Lp(a) PRS] | 2,083/2,736,287 | 0.76 (0.73, 0.79) | Reference |
| **IS** |  |  |  |
| 0-2 risk factor within target ranges | 21/9,800 | 2.14 (1.36, 2.90) | 1.03 (0.27, 1.78) |
| 3-4 risk factor within target ranges | 209/122,363 | 1.71 (1.48, 1.88) | 0.60 (0.37, 0.77) |
| 5-6 risk factor within target ranges | 407/344,049 | 1.18 (1.07, 1.28) | 0.07 (-0.03, 0.18) |
| 7-8 risk factor within target ranges | 135/206,640 | 0.65 (0.57, 0.75) | -0.46 (-0.56, -0.36) |
| Control [Q1-Q4 Lp(a) PRS] | 3,037/2,732,675 | 1.11 (1.07, 1.15) | Reference |
| **CVD death** |  |  |  |
| 0-2 risk factor within target ranges | 37/9,822 | 3.78 (2.73, 5.34) | 2.33 (1.31, 3.89) |
| 3-4 risk factor within target ranges | 339/122,403 | 2.77 (2.50, 3.04) | 1.33 (1.07, 1.60) |
| 5-6 risk factor within target ranges | 511/343,424 | 1.49 (1.40, 1.61) | 0.05 (-0.04, 0.19) |
| 7-8 risk factor within target ranges | 157/205,644 | 0.76 (0.67, 0.87) | -0.67 (-0.78, -0.55) |
| Control [Q1-Q4 Lp(a) PRS] | 3,915/2,725,201 | 1.44 (1.39, 1.47) | Reference |

Abbreviations: ASCVD, arteriosclerotic cardiovascular disease; CVD, cardiovascular disease; CI, confidence interval; HR, hazard ratio; Lp(a), lipoprotein(a); MI, myocardial infarction; PAD, peripheral arterial disease; PRS, polygenetic risk score; IS, ischemic stroke.

**Supplementary Table S15.** Incidence rates (per 1,000 person-y) for individual ASCVD outcomes according to the number of risk factor within target ranges among participants with elevated circulating Lp(a) compared with those with non-elevated circulating Lp(a) in ARIC.

|  | **Events/person-y** | **Incidence rate (95% CI)** | **Absolute rate difference (95% CI)** |
| --- | --- | --- | --- |
| **MI** |  |  |  |
| 0-3 risk factor within target ranges | 109/7,408 | 13.80 (11.50, 16.11) | 7.97 (5.52, 10.19) |
| 4-5 risk factor within target ranges | 212/26,435 | 7.82 (6.84, 8.77) | 1.94 (1.07, 2.96) |
| 6-7 risk factor within target ranges | 54/15,893 | 3.34 (2.38, 4.13) | -2.51 (-3.52, -1.67) |
| Control [Q1-Q4 Lp(a)] | 1,238/206,282 | 5.85 (5.57, 6.19) | Reference |
| **PAD** |  |  |  |
| 0-3 risk factor within target ranges | 40/8,929 | 4.48 (3.50, 5.71) | 3.00 (1.93, 4.20) |
| 4-5 risk factor within target ranges | 52/30,547 | 1.71 (1.32, 2.23) | 0.23 (-2.10, 0.74) |
| 6-7 risk factor within target ranges | 12/17,644 | 0.68 (0.33, 1.15) | -0.80 (-1.26, -0.35) |
| Control [Q1-Q4 Lp(a)] | 344/233,047 | 1.48 (1.36, 1.63) | Reference |
| **IS** |  |  |  |
| 0-3 risk factor within target ranges | 68/8,143 | 8.35 (6.49, 10.36) | 5.13 (3.17, 7.10) |
| 4-5 risk factor within target ranges | 97/28,130 | 3.45 (2.79, 4.06) | 0.23 (-0.45, 0.90) |
| 6-7 risk factor within target ranges | 43/16,332 | 2.63 (1.80, 3.31) | -0.59 (-1.38, 0.12) |
| Control [Q1-Q4 Lp(a)] | 695/215,670 | 3.22 (2.98, 3.44) | Reference |
| **CVD death** |  |  |  |
| 0-3 risk factor within target ranges | 92/8,671 | 10.60 (8.86, 12.91) | 7.4 (4.9, 10.2) |
| 4-5 risk factor within target ranges | 157/28,971 | 5.42 (4.68, 6.41) | 1.3 (0.3, 2.3) |
| 6-7 risk factor within target ranges | 42/16,661 | 2.52 (1.71, 3.37) | -2.1 (-2.8, -1.5) |
| Control [Q1-Q4 Lp(a)] | 1,070/221,983 | 4.82 (4.57, 5.10) | Reference |

Abbreviations: ASCVD, arteriosclerotic cardiovascular disease; ARIC, Atherosclerosis Risk In Communities; CVD, cardiovascular disease; CI, confidence interval; HR, hazard ratio; Lp(a), lipoprotein(a); MI, myocardial infarction; PAD, peripheral arterial disease; IS, ischemic stroke.

**Supplementary Table S16.** Age- and sex-specific HRs for ASCVD according to the number of risk factor within target ranges among participants with elevated circulating Lp(a) compared with those with non-elevated circulating Lp(a) in UK Biobank.

| ***P*-interaction = 0.573** | **Age stratification** | | | | | |
| --- | --- | --- | --- | --- | --- | --- |
|  | **<50 y** | | **50-59 y** | | **≥60 y** | |
|  | **Events/person-y** | **HR (95% CI)** | **Events/person-y** | **HR (95% CI)** | **Events/person-y** | **HR (95% CI)** |
| 0-2 risk factor within target ranges | 15/ 2370 | 2.91 (1.75, 4.85) | 47/4205 | 2.45 (1.83, 3.26) | 64/3232 | 2.23 (1.75, 2.86) |
| 3-4 risk factor within target ranges | 144/27303 | 2.59 (2.17, 3.08) | 387/53022 | 1.77 (1.59, 1.96) | 589/45483 | 1.59 (1.46, 1.73) |
| 5-6 risk factor within target ranges | 184/89047 | 1.23 (1.05, 1.44) | 574/143123 | 1.12 (1.03, 1.23) | 1153/128064 | 1.18 (1.11, 1.26) |
| 7-8 risk factor within target ranges | 50/70378 | 0.55 (0.42, 0.73) | 171/81333 | 0.68 (0.59, 0.80) | 347/57094 | 0.86 (0.77, 0.96) |
| Control [Q1-Q4 circulating Lp(a)] | 1,143/ 729678 | Reference | 3,985/1125103 | Reference | 7,464/974735 | Reference |
| ***P*-interaction = 0.400** | **Sex stratification** | | | | **-** | |
|  | **Women** | | **Men** | | **-** | |
|  | **Events/person-y** | **HR (95% CI)** | **Events/person-y** | **HR (95% CI)** | **-** | **-** |
| 0-2 risk factor within target ranges | 243/4616 | 2.31 (1.69, 3.17) | 87/5191 | 2.40 (1.94, 2.96) | **-** | **-** |
| 3-4 risk factor within target ranges | 683/63234 | 1.84 (1.67, 2.04) | 714/62573 | 1.68 (1.55, 1.81) | **-** | **-** |
| 5-6 risk factor within target ranges | 406/201635 | 1.10 (1.02, 1.20) | 1,228/158600 | 1.21 (1.14, 1.29) | **-** | **-** |
| 7-8 risk factor within target ranges | 39/133754 | 0.77 (0.68, 0.88) | 325/75052 | 0.76 (0.68, 0.84) | **-** | **-** |
| Control [Q1-Q4 circulating Lp(a)] | 4,748/1620461 | Reference | 7,844/1209056 | Reference | **-** | **-** |

The original model was adjusted for age, sex, ethnicity, TDI, FHx of CVD, antihyperglycemic drug use, antihypertensive drug use, and statin use. For sex stratification, the stratification factor was not adjusted for.

Abbreviations: ASCVD, arteriosclerotic cardiovascular disease; CI, confidence interval; HR, hazard ratio; Lp(a), lipoprotein(a).

**Supplementary Table S17.** Age- and sex-specific HRs for ASCVD according to the number of risk factor within target ranges among participants with high Lp(a) PRS compared with those with low Lp(a) PRS in UK Biobank.

| ***P*-interaction = 0.731** | **Age stratification** | | | | | |
| --- | --- | --- | --- | --- | --- | --- |
|  | **<50 y** | | **50-59 y** | | **≥60 y** | |
|  | **Events/person-y** | **HR (95% CI)** | **Events/person-y** | **HR (95% CI)** | **Events/person-y** | **HR (95% CI)** |
| 0-2 risk factor within target ranges | 13/2213 | 2.31 (1.34, 4.01) | 51/4101 | 2.76 (2.09, 3.63) | 58/3039 | 2.03 (1.57, 2.64) |
| 3-4 risk factor within target ranges | 131/25651 | 2.40 (2.00, 2.88) | 391/50524 | 1.78 (1.60, 1.98) | 546/42314 | 1.52 (1.39, 1.66) |
| 5-6 risk factor within target ranges | 170/84834 | 1.21 (1.03, 1.42) | 530/132290 | 1.10 (1.00, 1.20) | 1,081/119677 | 1.15 (1.08, 1.23) |
| 7-8 risk factor within target ranges | 48/71931 | 0.54 (0.40, 0.72) | 159/77185 | 0.67 (0.57, 0.79) | 326/54707 | 0.82 (0.74, 0.92) |
| Control [Q1-Q4 Lp(a) PRS] | 1,017/654889 | Reference | 3,767/1073996 | Reference | 7,306/954271 | Reference |
| ***P*-interaction = 0.531** | **Sex stratification** | | | | **-** | |
|  | **Women** | | **Men** | | **-** | |
|  | **Events/person-y** | **HR (95% CI)** | **Events/person-y** | **HR (95% CI)** | **-** | **-** |
| 0-2 risk factor within target ranges | 35/3983 | 2.45 (1.75, 3.42) | 87/5370 | 2.28 (1.85, 2.82) | - | - |
| 3-4 risk factor within target ranges | 342/54941 | 1.76 (1.58, 1.96) | 726/63550 | 1.64 (1.52, 1.76) | - | - |
| 5-6 risk factor within target ranges | 591/179418 | 1.08 (0.99, 1.18) | 1,190/157383 | 1.17 (1.10, 1.25) | - | - |
| 7-8 risk factor within target ranges | 223/128982 | 0.76 (0.66, 0.87) | 310/74842 | 0.72 (0.64, 0.81) | - | - |
| Control [Q1-Q4 Lp(a) PRS] | 4,680/1553588 | Reference | 7,410/1129569 | Reference | - | - |

The original model was adjusted for age, sex, ethnicity, TDI, FHx of CVD, antihyperglycemic drug use, antihypertensive drug use, statin use, and the first 10 principal components of ancestry. For sex stratification, the stratification factor was not adjusted for.

Abbreviations: ASCVD, arteriosclerotic cardiovascular disease; CI, confidence interval; HR, hazard ratio; Lp(a), lipoprotein(a); PRS, polygenetic risk score.

**Supplementary Table S18.** Age- and sex-specific HRs for ASCVD according to the number of risk factor within target ranges among participants with elevated circulating Lp(a) compared with those with non-elevated circulating Lp(a) in ARIC.

| ***P*-interaction = 0.769** | **Age stratification** | | | | | |
| --- | --- | --- | --- | --- | --- | --- |
|  | **<50 y** | | **50-59 y** | | **≥60 y** | |
|  | **Events/person-y** | **HR (95% CI)** | **Events/person-y** | **HR (95% CI)** | **Events/person-y** | **HR (95% CI)** |
| 0-3 risk factor within target ranges | 34/1,944 | 2.11 (1.48, 3.01) | 118/4,029 | 2.05 (1.69, 2.48) | 52/1,434 | 1.76 (1.31, 2.36) |
| 3-5 risk factor within target ranges | 73/8,073 | 1.37 (1.07, 1.76) | 204/14,736 | 1.13 (0.97, 1.31) | 97/3,625 | 1.37 (1.10, 1.69) |
| 6-7 risk factor within target ranges | 20/4,691 | 0.78 (0.50, 1.23) | 77/9,138 | 0.80 (0.63, 1.00) | 25/2,063 | 0.65 (0.43, 0.97) |
| Control [Q1-Q4 circulating Lp(a)] | 445/64,593 | Reference | 1,407/112,321 | Reference | 605/29,367 | Reference |
| ***P*-interaction = 0.260** | **Sex stratification** | | | | **-** | |
|  | **Women** | | **Men** | | **-** | |
|  | **Events/person-y** | **HR (95% CI)** | **Events/person-y** | **HR (95% CI)** | **-** | **-** |
| 0-3 risk factor within target ranges | 89/2,860 | 2.12 (1.69, 2.63) | 115/4,548 | 1.77 (1.45, 2.16) | - | - |
| 3-5 risk factor within target ranges | 184/9,800 | 1.30 (1.11, 1.52) | 190/16,635 | 1.16 (0.99, 1.35) | - | - |
| 6-7 risk factor within target ranges | 55/5,370 | 0.78 (0.60, 1.03) | 67/10,523 | 0.74 (0.58, 0.95) | - | - |
| Control [Q1-Q4 circulating Lp(a)] | 1,329/90,849 | Reference | 1,128/115,432 | Reference | - | - |

The original model was adjusted for age, sex, ethnicity, household income, FHx of CVD, antihyperglycemic drug use, antihypertensive drug use, and statin use.. For sex stratification, the stratification factor was not adjusted for.

Abbreviations: ASCVD, arteriosclerotic cardiovascular disease; CI, confidence interval; HR, hazard ratio; Lp(a), lipoprotein(a)

**Supplementary Table S19.** HRs for ASCVD according to the number of risk factor within target ranges among participants with elevated Lp(a) compared with those with non-elevated Lp(a) in UK Biobank and ARIC, stratified by 10-y ASCVD risk.

| ***P*-interaction < 0.001** | **10-y ASCVD risk stratification**  **Circulating Lp(a) in UK Biobank** | | | |
| --- | --- | --- | --- | --- |
|  | **QRISK3 <10%** | | **QRISK3 ≥10%** | |
|  | **Events/person-y** | **HR (95% CI)** | **Events/person-y** | **HR (95% CI)** |
| 0-2 risk factor within target ranges | 17/2,497 | 3.09 (1.92, 4.97) | 109/7,328 | 2.03 (1.68, 2.45) |
| 3-4 risk factor within target ranges | 246/47,517 | 2.13 (1.87, 2.43) | 874/78,290 | 1.53 (1.43, 1.64) |
| 5-6 risk factor within target ranges | 561/187,196 | 1.33 (1.22, 1.46) | 1,350/173,038 | 1.11 (1.05, 1.18) |
| 7-8 risk factor within target ranges | 238/148,344 | 0.84 (0.74, 0.96) | 330/60,462 | 0.81 (0.73, 0.90) |
| Control [Q1-Q4 circulating Lp(a)] | 3,277/1,532,900 | Reference | 9,315/1,296,617 | Reference |
| ***P*-interaction = 0.002** | **10-y ASCVD risk stratification**  **Lp(a) PRS in UK Biobank** | | | |
|  | **QRISK3 <10%** | | **QRISK3 ≥10%** | |
|  | **Events/person-y** | **HR (95% CI)** | **Events/person-y** | **HR (95% CI)** |
| 0-2 risk factor within target ranges | 16/2,339 | 2.98 (1.82, 4.87) | 106/7,015 | 1.99 (1.48, 1.59) |
| 3-4 risk factor within target ranges | 238/45,015 | 2.10 (1.84, 2.39) | 830/73,476 | 1.48 (1.38, 1.59) |
| 5-6 risk factor within target ranges | 525/179,206 | 1.26 (1.15, 1.39) | 1,256/157,596 | 1.09 (1.03, 1.16) |
| 7-8 risk factor within target ranges | 220/147,468 | 0.79 (0.69, 0.90) | 313/56,356 | 0.80 (0.72, 0.90) |
| Control [Q1-Q4 Lp(a) PRS] | 3,077/1,421,095 | Reference | 9,013/1,262,062 | Reference |
| ***P*-interaction = 0.097** | **10-y ASCVD risk stratification**  **Circulating Lp(a) in ARIC** | | | |
|  | **PCE <7.5%** | | **PCE ≥7.5%** | |
|  | **Events/person-y** | **HR (95% CI)** | **Events/person-y** | **HR (95% CI)** |
| 0-3 risk factor within target ranges | 48/2,812 | 2.22 (1.66, 2.98) | 156/4,596 | 1.47 (1.24, 1.74) |
| 3-5 risk factor within target ranges | 202/20,253 | 1.33 (1.14, 1.55) | 172/6,181 | 1.21 (1.03, 1.42) |
| 6-7 risk factor within target ranges | 91/14,110 | 0.88 (0.71, 1.09) | 31/1,782 | 0.74 (0.52, 1.06) |
| Control [Q1-Q4 circulating Lp(a)] | 1,112/148,496 | Reference | 1,345/57,785 | Reference |

The original model was adjusted for age, sex, ethnicity, household income, FHx of CVD, antihyperglycemic drug use, antihypertensive drug use, and statin use.

Abbreviations: ASCVD, arteriosclerotic cardiovascular disease; CI, confidence interval; HR, hazard ratio; Lp(a), lipoprotein(a)

**Supplementary Table S20.** Sensitivity analyses for the HRs for ASCVD according to the number of risk factor within target ranges among participants with elevated circulating Lp(a) compared with those with non-elevated circulating Lp(a) in UK Biobank.

| **Control was participants with circulating Lp(a) <120 nmol/L (≈50 mg/dL)** | **Events/person-y** | **HR (95% CI)** |
| --- | --- | --- |
| 0-2 risk factor within target ranges | 78/5,980 | 2.65 (2.12, 3.31) |
| 3-4 risk factor within target ranges | 704/78,202 | 1.76 (1.63, 1.90) |
| 5-6 risk factor within target ranges | 1200/217,761 | 1.21 (1.14, 1.28) |
| 7-8 risk factor within target ranges | 351/120,347 | 0.79 (0.71, 0.87) |
| Control [Circulating Lp(a) <120 nmol/L (≈50 mg/dL)] | 13,984/3,111,884 | Reference |
| **Control was participants with non-elevated Lp(a) and 4 risk factor within target ranges** | **Events/person-y** | **HR (95% CI)** |
| 0-2 risk factor within target ranges | 126/9,807 | 1.82 (1.52, 2.18) |
| 3-4 risk factor within target ranges | 1120/125,808 | 1.31 (1.22, 1.40) |
| 5-6 risk factor within target ranges | 1911/360,235 | 0.87 (0.82, 0.92) |
| 7-8 risk factor within target ranges | 568/208,806 | 0.55 (0.50, 0.60) |
| Control [Q1-Q4 Lp(a) & 4 risk factor within target ranges] | 2,498/357,808 | Reference |
| **Control was participants with lowest quintile of circulating Lp(a)** | **Events/person-y** | **HR (95% CI)** |
| 0-2 risk factor within target ranges | 126/9807 | 2.53 (2.12, 3.02) |
| 3-4 risk factor within target ranges | 1,120/125808 | 1.83 (1.71, 1.96) |
| 5-6 risk factor within target ranges | 1,911/360235 | 1.23 (1.16, 1.30) |
| 7-8 risk factor within target ranges | 568/208806 | 0.79 (0.72, 0.86) |
| Control [Q1 Lp(a)] | 3,046/710,724 | Reference |
| **Exclusion of participants who developed ASCVD within 5 years of follow-up** | **Events/person-y** | **HR (95% CI)** |
| 0-2 risk factor within target ranges | 87/9,645 | 2.24 (1.82, 2.77) |
| 3-4 risk factor within target ranges | 837/124,566 | 1.74 (1.63, 1.87) |
| 5-6 risk factor within target ranges | 1,443/357,879 | 1.18 (1.12, 1.25) |
| 7-8 risk factor within target ranges | 442/208,025 | 0.78 (0.70, 0.85) |
| Control [Q1-Q4 Lp(a)] | 9,492/2,812,751 | Reference |

The model was adjusted for age, sex, ethnicity, TDI, FHx of CVD, antihyperglycemic drug use, antihypertensive drug use, and statin use.

Abbreviations: ap(B), apolipoprotein B; ASCVD, arteriosclerotic cardiovascular disease; CI, confidence interval; HR, hazard ratio; Lp(a), lipoprotein(a)

**Supplementary Table S21.** Sensitivity analyses for the HRs for ASCVD according to the number of risk factor within target ranges among participants with high Lp(a) PRS compared with those with low Lp(a) PRS in UK Biobank.

| **Control was participants with low Lp(a) PRS and 4 risk factor within target ranges** | **Events/person-y** | **HR (95% CI)** |
| --- | --- | --- |
| 0-2 risk factor within target ranges | 122/9,354 | 1.78 (1.47, 2.14) |
| 3-4 risk factor within target ranges | 1,068/118,491 | 1.27 (1.18, 1.37) |
| 5-6 risk factor within target ranges | 1,781/336,801 | 0.84 (0.79, 0.90) |
| 7-8 risk factor within target ranges | 533/203,824 | 0.53 (0.48, 0.58) |
| Control [Q1-Q4 Lp(a) & 4 risk factor within target ranges] | 2,399/341,613 | Reference |
| **Control was participants with lowest quintile of Lp(a) PRS** | **Events/person-y** | **HR (95% CI)** |
| 0-2 risk factor within target ranges | 122/9,354 | 2.45 (2.04, 2.93) |
| 3-4 risk factor within target ranges | 1,068/118,491 | 1.76 (1.64, 1.89) |
| 5-6 risk factor within target ranges | 1,781/336,801 | 1.19 (1.12, 1.26) |
| 7-8 risk factor within target ranges | 533/203,824 | 0.75 (0.69, 0.83) |
| Control [Q1 Lp(a)] | 2,888/671,279 | Reference |
| **Exclusion of participants who developed ASCVD within 5 years of follow-up** | **Events/person-y** | **HR (95% CI)** |
| 0-2 risk factor within target ranges | 90/9,212 | 2.37 (1.93, 2.92) |
| 3-4 risk factor within target ranges | 794/117,328 | 1.69 (1.57, 1.82) |
| 5-6 risk factor within target ranges | 1,359/334,570 | 1.16 (1.10, 1.23) |
| 7-8 risk factor within target ranges | 422/203,086 | 0.77 (0.69, 0.84) |
| Control [Q1-Q4 Lp(a)] | 9,078/2,667,009 | Reference |

The model was adjusted for age, sex, ethnicity, TDI, FHx of CVD, antihyperglycemic drug use, antihypertensive drug use, statin use, and the first 10 principal components of ancestry.

Abbreviations: ap(B), apolipoprotein B; ASCVD, arteriosclerotic cardiovascular disease; CI, confidence interval; HR, hazard ratio; Lp(a), lipoprotein(a); PRS, polygenetic risk score.

**Supplementary Table S22.** Sensitivity analyses for the HRs for ASCVD according to the number of risk factor within target ranges among participants with elevated circulating Lp(a) compared with those with non-elevated circulating Lp(a) in ARIC.

| **Control was participants with circulating <Lp(a) 50 mg/dL** | **Events/person-y** | **HR (95% CI)** |
| --- | --- | --- |
| 0-3 risk factor within target ranges | 246/84,09 | 1.96 (1.71, 2.26) |
| 4-5 risk factor within target ranges | 384/27,438 | 1.15 (1.03, 1.29) |
| 6-7 risk factor within target ranges | 119/14,378 | 0.78 (0.65, 0.93) |
| Control [Q1-Q4 Lp(a) & 3-4 risk factor within target ranges] | 2,408/205,793 | Reference |
| **Control was participants with non-elevated circulating Lp(a) and 3-4 risk factor within target ranges** | **Events/person-y** | **HR (95% CI)** |
| 0-3 risk factor within target ranges | 204/7,408 | 2.12 (1.82, 2.46) |
| 4-5 risk factor within target ranges | 374/26,435 | 1.28 (1.14, 1.44) |
| 6-7 risk factor within target ranges | 122/15,893 | 0.77 (0.64, 0.93) |
| Control [Q1-Q4 Lp(a) & 3-4 risk factor within target ranges] | 1,316/113,341 | Reference |
| **Control was participants with lowest quintile of circulating Lp(a)** | **Events/person-y** | **HR (95% CI)** |
| 0-3 risk factor within target ranges | 204/7,408 | 2.02 (1.72, 2.38) |
| 4-5 risk factor within target ranges | 374/26,435 | 1.23 (1.08, 1.40) |
| 6-7 risk factor within target ranges | 122/15,893 | 0.76 (0.62, 0.93) |
| Control [Q1 Lp(a)] | 637/52,780 | Reference |
| **Exclusion of participants who developed ASCVD within 5 years of follow-up** | **Events/person-y** | **HR (95% CI)** |
| 0-3 risk factor within target ranges | 117/7,299 | 1.94 (1.64, 2.29) |
| 4-5 risk factor within target ranges | 343/26,270 | 1.25 (1.11, 1.40) |
| 6-7 risk factor within target ranges | 113/15,846 | 0.76 (0.63, 0.92) |
| Control [Q1-Q4 Lp(a)] | 2,221/205,095 | Reference |

The model was adjusted for age, sex, ethnicity, family income, FHx of CVD, antihyperglycemic drug use, antihypertensive drug use, and statin use.

Abbreviations: ASCVD, arteriosclerotic cardiovascular disease; CI, confidence interval; HR, hazard ratio; Lp(a), lipoprotein(a)

### Supplementary Figure S1. Flowchart of participant enrolment.

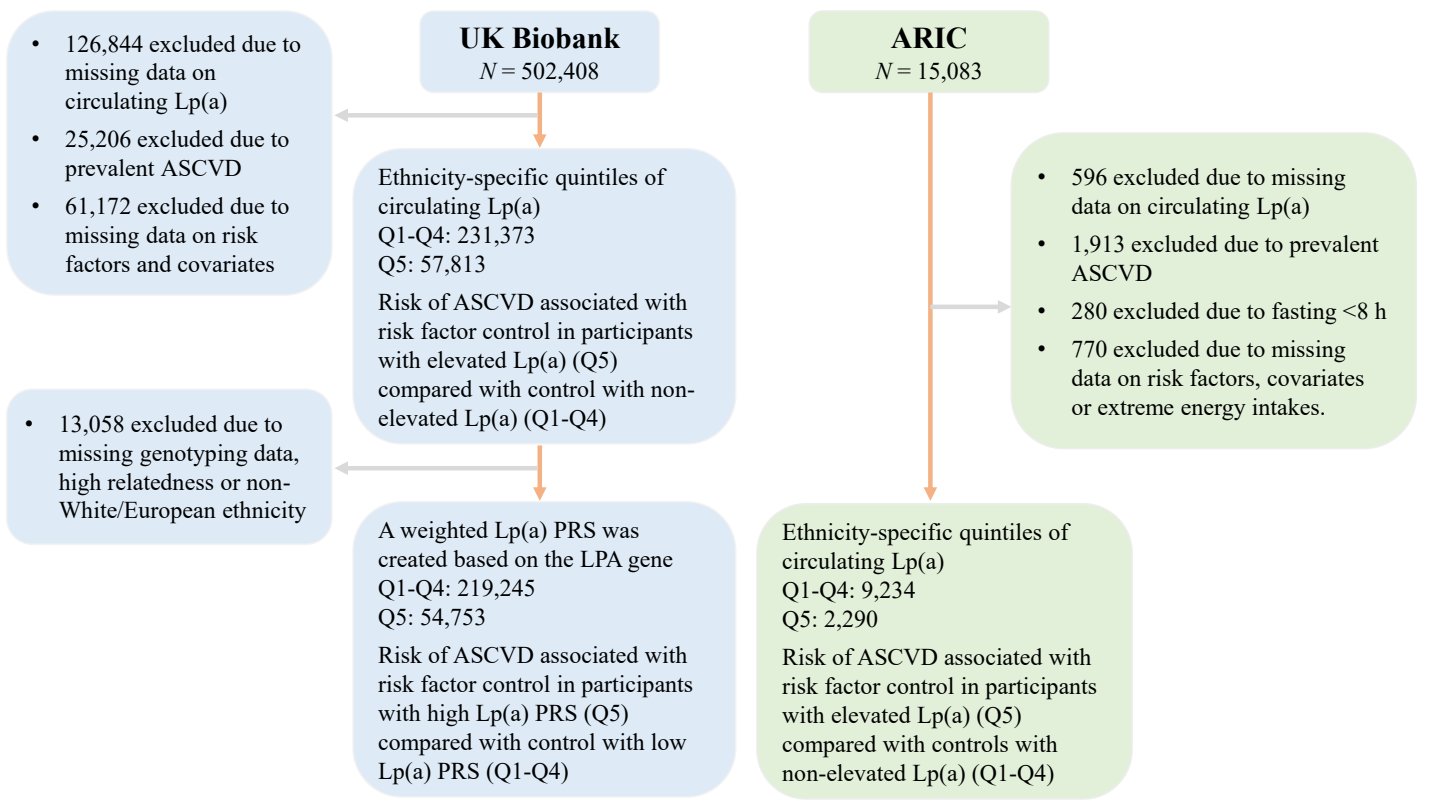

ASCVD, atherosclerotic cardiovascular disease; ARIC, Atherosclerosis Risk in Communities; Lp(a), lipoprotein(a); PRS, polygenetic risk score.

**Supplementary Figure S2.** Relative importance of risk factors for predicting individual ASCVD outcomes among participants with elevated circulating Lp(a) in UK Biobank.

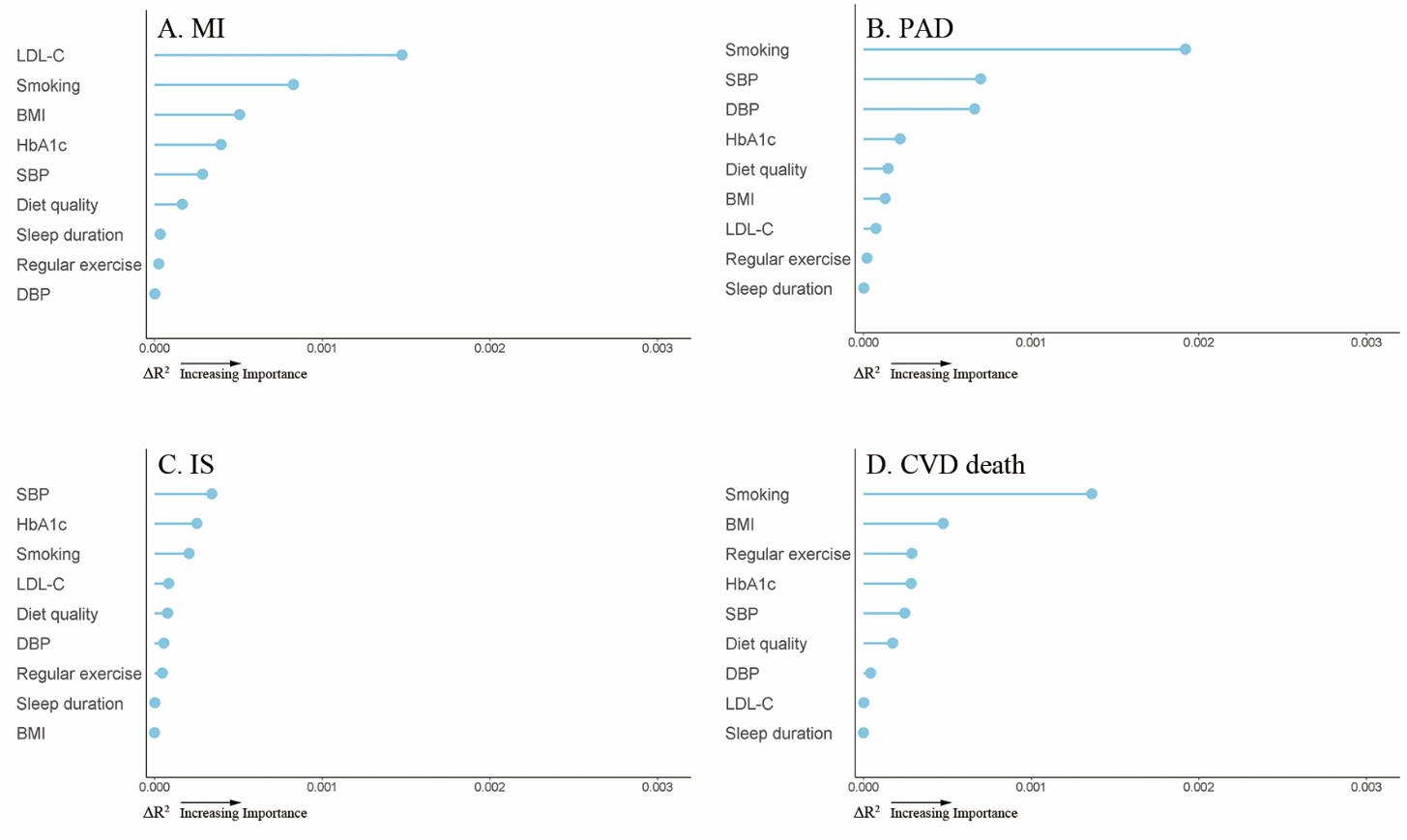

The ΔR^2^ value for each risk factor was computed by subtracting the R^2^ value of the Cox model that did not include the specific risk factor from the R^2^ value generated by the Cox model that included all the risk factors.

BMI, body mass index; CVD, cardiovascular disease; DBP, diastolic blood pressure; HbA1c, glycosylated hemoglobin; IS, ischemic stroke; LDL-C, low density lipoprotein cholesterol; MI, myocardial infarction; PAD, peripheral arterial disease; SBP, systolic blood pressure

**Supplementary Figure S3.** Relative importance of risk factors for predicting individual ASCVD outcomes among participants with high Lp(a) PRS in UK Biobank.

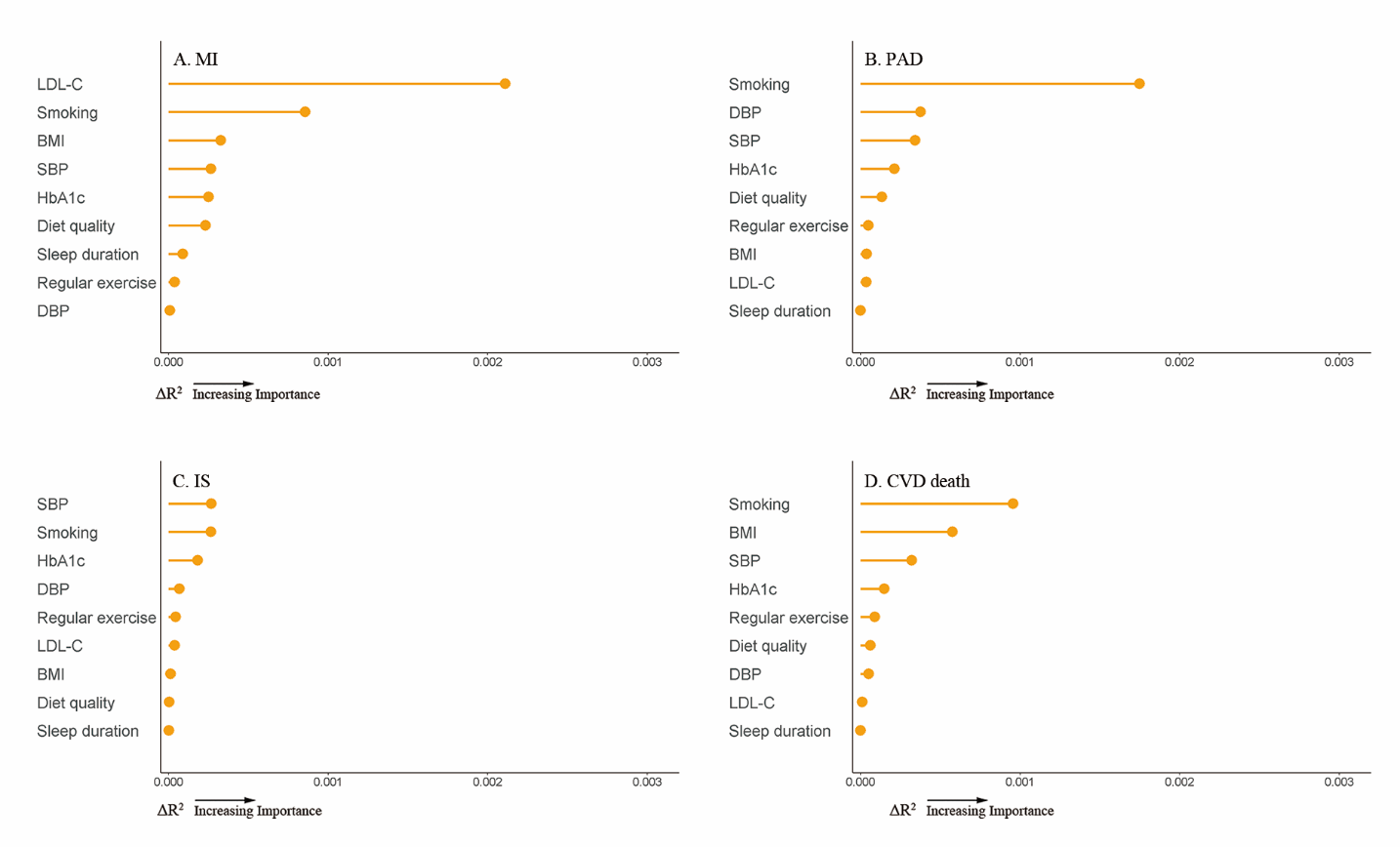

All analyses were conducted among the White. The ΔR^2^ value for each risk factor was computed by subtracting the R^2^ value of the Cox model that did not include the specific risk factor from the R^2^ value generated by the Cox model that included all the risk factors plus the first 10 principal components of ancestry.

BMI, body mass index; CVD, cardiovascular disease; DBP, diastolic blood pressure; HbA1c, glycosylated hemoglobin; IS, ischemic stroke; LDL-C, low density lipoprotein cholesterol; MI, myocardial infarction; PAD, peripheral arterial disease; SBP, systolic blood pressure

**Supplementary Figure S4.** Relative importance of risk factors for predicting individual ASCVD outcomes among participants with elevated circulating Lp(a) in ARIC.

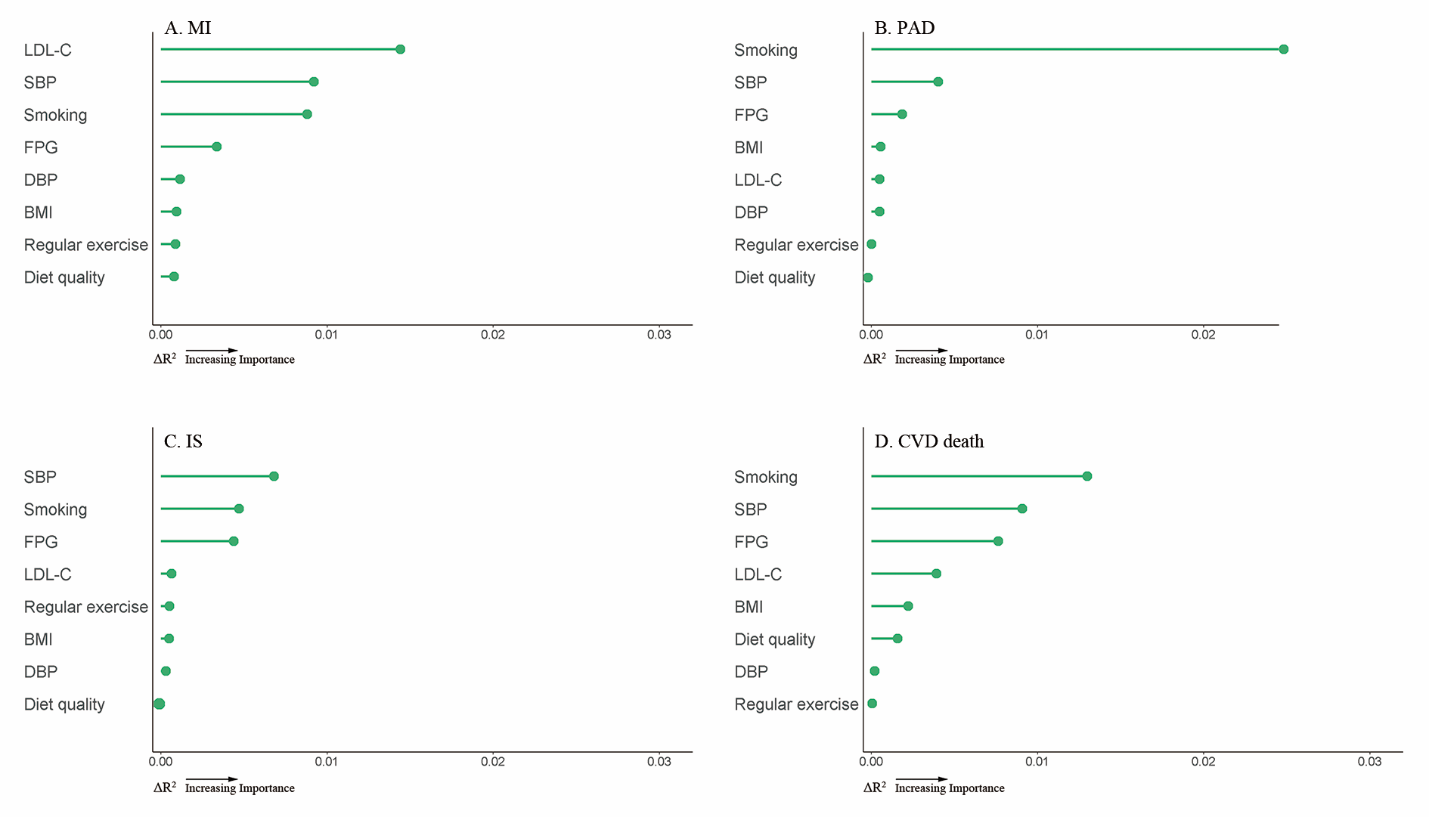

The ΔR^2^ value for each risk factor was computed by subtracting the R^2^ value of the Cox model that did not include the specific risk factor from the R^2^ value generated by the Cox model that included all the risk factors.

BMI, body mass index; CVD, cardiovascular disease; DBP, diastolic blood pressure; FPG, fasting plasma glucose; HbA1c, glycosylated hemoglobin; LDL-C, low density lipoprotein cholesterol; MI, myocardial infarction; PAD, peripheral arterial disease; IS, ischemic stroke.
